# The Inter-Connectedness of Post-Traumatic Stress Disorder (PTSD) Symptomatology Across the Lifespan

**DOI:** 10.64898/2026.01.25.26344779

**Authors:** David Johnston, Nimrod Hertz-Palmor, Caitlin Hitchcock, Melissa Black, Anke de Haan, Richard Bryant, Anna McKinnon, Alexandra De Young, Patrick Smith, Richard Meiser-Stedman, Anna Bevan, Tim Dalgleish

## Abstract

PTSD is defined by a core of inter-connected symptom clusters. It is currently unclear whether this pattern of interconnections remains stable across the lifespan or differs across key developmental periods. Synthesising seven international trauma-exposed samples (N=5,470), we compared network interrelationships among core self-reported and/or caregiver-reported PTSD symptom clusters (re-experiencing, avoidance and arousal) in preschoolers, school-aged children, adolescents, and adults. For self-report from school-age to adulthood, within-cluster connectivity (associations among symptoms within the same cluster) was consistently stronger than between-cluster connectivity (associations among symptoms from different clusters) across all age groups. This was especially true for the re-experiencing symptom cluster. These inter-relationships appeared stable across with lifecourse with no significant age-related differences. In contrast, caregiver reports from preschool to adolescence, showed stronger within-cluster connectivity among arousal symptoms but these only emerged in school-aged children and adolescents. Longitudinal analyses across approximately the first year post-trauma of self-report symptoms indicated that adults’ overall symptom connectivity increased over time, whereas school-aged children’s and adolescents’ networks became sparser. These findings suggest that while PTSD network architecture is broadly stable across different age groups, reporter discrepancies and temporal dynamics warrant close attention, especially with regards to less visible, internalised symptoms such as re-experiencing.

---

Psychological distress following the experience of a traumatic event has been recognised throughout history. In 1980 this constellation of post-trauma psychological symptoms was formally characterised as Post-traumatic Stress Disorder (PTSD) in the Third Edition of the Diagnostic and Statistical Manual for Mental Disorders (DSM-III)^1^. PTSD is unusual among DSM syndromes in that the diagnostic criteria specify an aetiologic event, and that the symptoms must begin or worsen after the event. The defining symptoms of PTSD have evolved extensively between the DSM-III and DSM-5 with an overall increase in the number of symptoms from 12 to 20 and in the number of symptom criteria (clusters of similar symptoms) from three to four^2^. The DSM-5 also introduced the first age-specific PTSD subtype for children 6 years and younger to reflect unique developmental differences in symptom expression.

Establishing the symptom structure of PTSD is important for understanding its aetiology and maintenance. Cognitive theories of PTSD share the assumption that the attempt to assimilate new information (the traumatic experience) within a pre-existing set of beliefs and models of the world shapes an individual’s post-traumatic reactions^3^. The symptoms which together comprise the post-traumatic reaction are many and varied, and it has long been debated which symptoms define the core prototypical PTSD response. Reflective of this are the contrasting approaches of the two major classification systems (the DSM and the International Classification of Diseases [ICD]). The ICD proposes a tighter diagnosis focused on three core symptom clusters (re-experiencing, avoidance and hyperarousal), with more complex cases involving additional clusters being subsumed under a separate diagnosis of Complex PTSD. In contrast, the DSM eschews this differentiation, and aggregates all presentations under a single more extensive PTSD diagnosis that includes a criterion capturing negative alterations in cognition and mood^4^. Revision of the symptom clusters and composition over time for the DSM and ICD has relied on guidance from factor analytic research in order to determine the most parsimonious algorithms^5^. The central tenet of Network Theory is that a focus on these clusters, and the symptoms they contain, in isolation, misses important information concerning the relationships between them^6^, and that a dynamic approach to symptom and cluster interrelations is key for truly understanding mental disorders including PTSD^7^ .

Network theory conceptualises mental disorders as a series of dynamic interactions between symptoms^6,8^. Within this framework, ‘node centrality’ reflects how strongly a given symptom or cluster is interconnected with others. This is important because central, more inter-connected nodes represent optimal first intervention targets, as any therapeutic gains have high potential to propagate across the associated symptoms. Understanding how such metrics of centrality differ for different trauma-exposed populations could therefore be critical to the endeavour to stratify interventions for maximum impact^8^. McNally et al.,^9^ pioneered this approach, finding support for an associative structure consistent with extant cognitive models of PTSD^10^, with hypervigilance playing a central role. Subsequent studies investigating different populations have found differences in which symptoms appear most central. For example in motor vehicle collision survivors, Bryant et al.,^11^ found reexperiencing symptoms (intrusions and physiological reactivity) to be most central, while in Ross et al.’s^12^ investigation in combat veterans, symptoms from Criteria D (negative alterations in mood and cognitions; negative emotional state and detachment) and E (hyperarousal; exaggerated startle response) were as central as Criterion B re-experiencing symptoms (nightmares and recurrent thoughts). Prompted by concerns about replicability and statistical power within the network field^6,13–16^, Fried et al^17^ investigated the structure of PTSD in four different trauma samples. While there were moderate to high correlations between the networks across the samples, with the most central symptoms consistently being intrusions, detachment, and hyper-reactivity, there were clear differences between the networks in terms of the associations between specific symptom pairs. This aligns with an increasing focus on how networks are modulated by individual differences with studies exploring how gender^18–20^, age ^21^, and trauma type^22^ influence the network symptom interrelationships. However, to date, most of this work has focused on adult trauma survivors.

The extant network analyses of PTSD symptoms in children and adolescents have tended to reflect findings in the adult literature^18,23–30^, with the predominant feature being the centrality of PTSD re-experiencing symptoms^11,31,32^. However, there are exceptions to this. One investigation of DSM-5 PTSD symptoms in school-aged children found trauma-related cognitions and persistent negative emotional state to be more central than reexperiencing symptomatology^23^. Additional studies found network differences between children and adults during COVID-19^28^, and between younger children and older adolescents exposed to disasters^29^ and war zones^30^. Most notably, older participants showed stronger global connectivity: adolescents had more strongly connected networks than children, and young adults showed stronger global connectivity than adolescents.

Local network properties, however, yielded mixed results in those age group comparisons, with some studies identifying arousal symptoms as central^28,29^, while others highlighted re-experiencing and avoidance^30^. Importantly, these studies focused on individual symptoms rather than examining symptom clusters, which may partly explain the inconsistency in findings. Yet, these differences suggest that posttraumatic stress symptoms (PTSS) are experienced differently in younger populations and that the nature of the inter-relationships may change across age, with consequent implications for how we should tailour interventions for different age groups^33,34^. The first aim of the present study was therefore to compare the network structures across different age groups across the lifespan from preschool to adulthood.

Another critical factor in elucidating PTSD symptom inter-relationships in children and adolescents is the influence of reporter identity. Unlike for adults, it is is often critical to obtain caregiver-report of symptoms, especially for youger children. Interestingly, substantial discrepancies have consistently been observed between caregiver-reported and child self-reported PTSD symptoms^35–40^. Such low parent-child agreement has been broadly linked to higher child symptom severity^39^, absence of a medical condition^36^, younger child age^37^, temporal proximity to the traumatic event,^37^, and greater parental PTSD symptomatology^35,38,40^. What we do not know is whether the patterns of symptom associations – the networks – also differ as a function of child vs caregiver report. The second aim of this study was therefore to to compare child self-report and caregiver-report network structures for preschool, school-age and adolescent samples.

Network analysis has also been used to investigate dynamic changes in PTSD symptom network structure in trauma-exposed populations across time. For example, in a longitudinal investigation in adults, Bryant et al^11^ found changes in symptom dynamics and overall network strength across time, which conformed to predictions from fear conditioning models of PTSD. Specifically, they revealed acute centrality and strengthening over time of re-experiencing and hyperarousal symptoms. These findings echo the centrality of current threat in theoretical models such as those of Ehlers and Clark^10^, and align with neurobiological accounts emphasizing hyperactivation of fear and threat-related neural circuits, particularly within the amygdala–hippocampal–prefrontal network^41^. A small number of other studies have investigated changes in the dynamic network structure of PTSD symptoms over time. Segal et al.,^42^ compared networks of combat soldiers 6 months pre- and post-deployment and found that, following deployment, symptoms of reactivity to triggers, and avoidance symptoms, became more closely associated. In a longitudinal study in children and adolescents measured at 2 weeks, 3 months and 6 months after a traumatic event, Ge et al.,^43^ found a strengthening of connectivity in the symptoms and the centrality of re-experiencing symptoms. However, we currently know little about how temporal changes differ as a function of age. This is important because knowing when to intervene and how in the weeks and months after a trauma is a crucial public health question and so elucidating any differences in the temporal evolution of symptom networks in this critical period will be critically informative. The final aim of the present study was therefore to compare temporal shifts in network structure between adults and youth in the immediate and subsequent aftermath of trauma.

The current study aimed to investigate differences in PTSD symptom associations and network structure across age groups. The overarching aims were to: (1) examine structural differences in self-report networks of children, adolescents, and adults; (2) compare caregiver reports for school-aged-children and adolescents aged six to eighteen years with those for preschool-aged children aged two to six years, for whom the DSM-5 specifies a distinct set of PTSD criteria reflecting developmental differences in symptom expression (e.g., play reenactment, fewer avoidance criteria, and greater reliance on behavioural manifestations)^44^; (3) to contrast networks of school-aged-children and adolescents when based on self-report versus caregiver report; and (4) to explore longitudinal changes in self-report network structure among school-aged-children and adolescents compared with adults. Unlike much of the network literature, which often yields unreplicable structures due to sample-specific patterns of symptom association, our analyses focused on symptom clusters rather than individual symptoms to capture broader patterns of association and connectivity beyond specific links and account for use of different measures.

Data were synthesised across seven samples across the UK, US and Australia (overall N=5,470, data-points=6,638). Samples 1 and 2 included caregiver reports on preschool-aged children (aged 2–6 years, Sample 1, N=520) and school-aged children and adolescents (aged 6.5–18 years, Sample 2, N=730). Samples 3–5 comprised self-reports from elementary-school-aged children (aged 6.5–12 years, Sample 3, N=1,079), adolescents (aged 13–18 years, Sample 4, N=978), and adults (Sample 5, N=995). Samples 6 and 7 contained longitudinal self-reported data from school-aged children and adolescents (Sample 6, N=359) and adults (Sample 7, N=809). All samples included individuals exposed to various types of trauma, including unintentional injuries, acute medical events, motor vehicle accidents, interpersonal violence, disasters, and work-related incidents, with partial overlap between caregiver- and self-reported data in children and adolescents. See Table 1 for Sample characteristics.

**Table 1.**
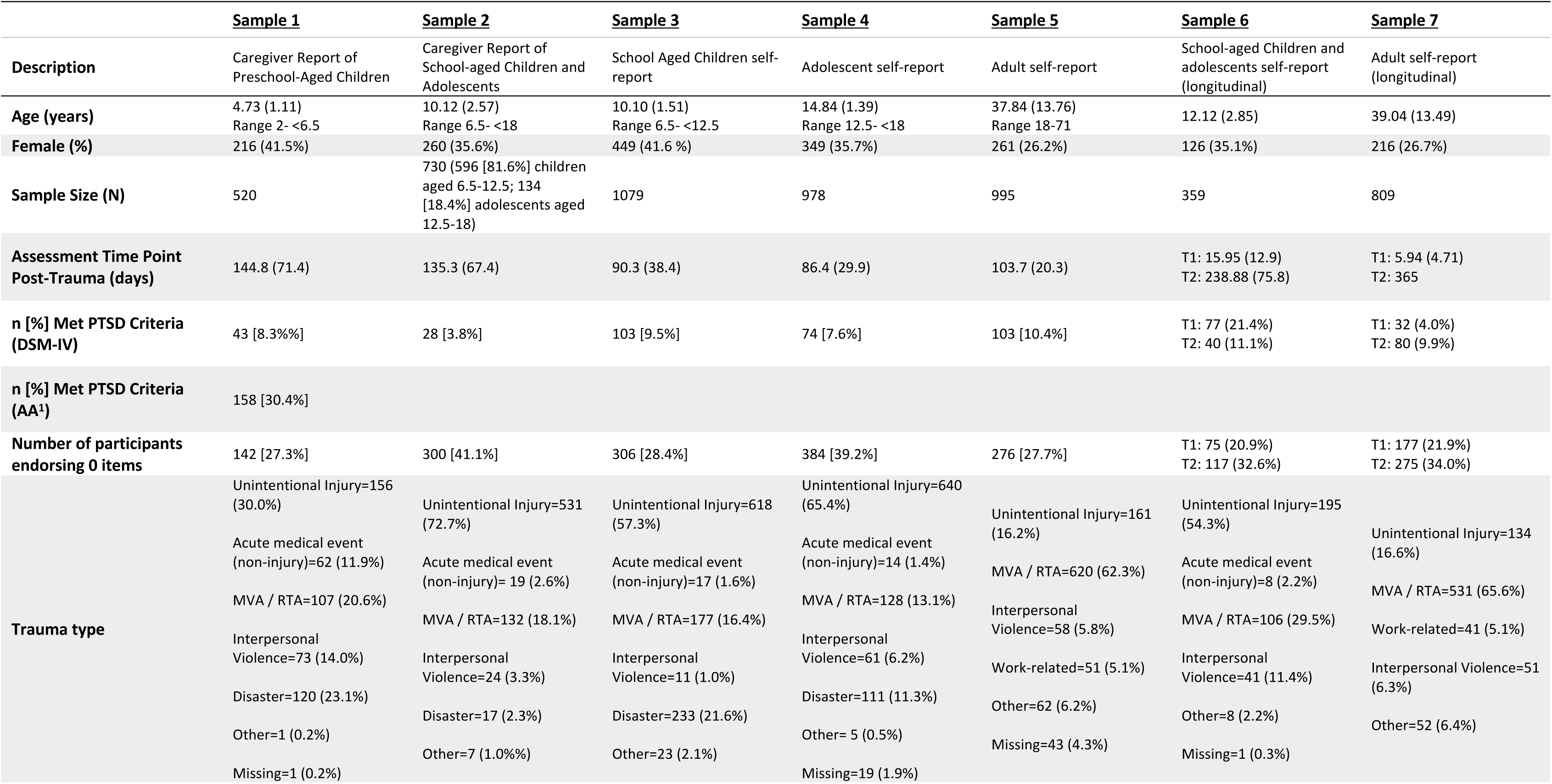
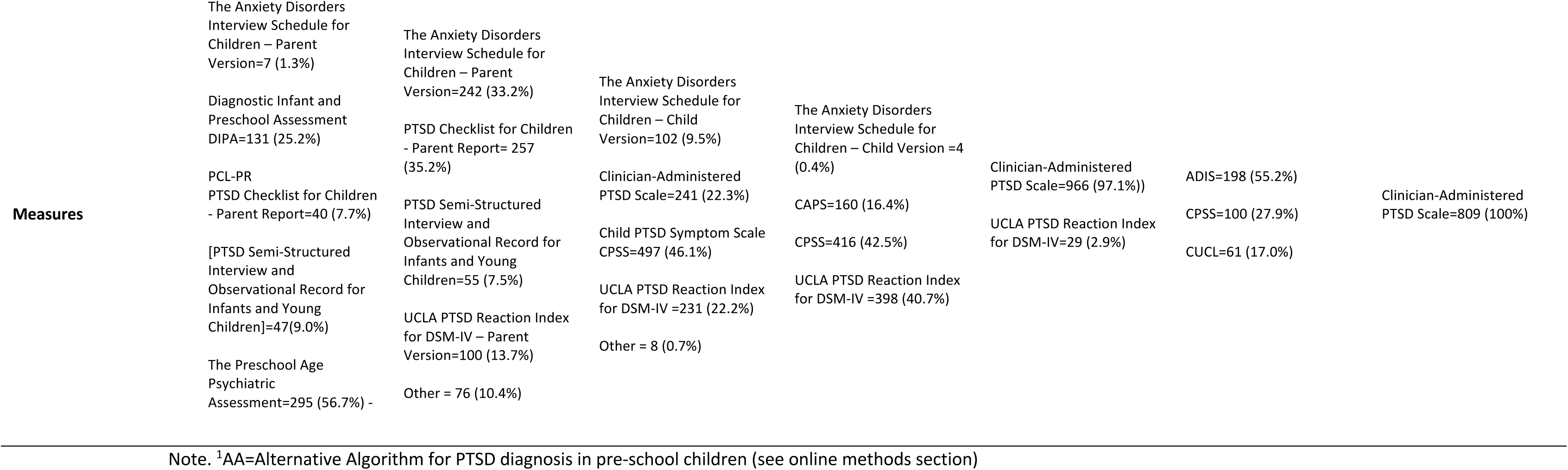
An overview of the characteristics of each sample. Data are mean (SD) unless otherwise specified.

Given the previously noted instability of individual symptom networks, we focused our analyses on associations within- and between-symptom clusters rather than just on individual symptoms. Our thesis is that cluster-level analyses are likely to capture broader, more stable patterns within the overall structure of the symptomatology, whereas analyses at the level of individual symptoms run the risk of overfitting in any given sample risking non-replicability. Moreover, focusing on symptom clusters that are largely consistent across DSM and ICD frameworks, enabled us to examine structures that better reflect the shared conceptualisation of PTSD. For the three PTSD clusters common to both DSM and ICD taxonomies^1^ —re-experiencing, avoidance, and arousal—we computed ‘cluster centrality’ (the overall influence of each clusters’ set of symptoms within the network) and ‘cluster connectivity’, which reflects the strength of associations either within symptom clusters (within-cluster connectivity) or between different clusters (between-cluster connectivity). We examined reporter identity effects by comparing self-and caregiver-reports for school-aged-children and adolescents (Samples 2–4) and age-related differences by comparing age groups in caregiver reports (Samples 1–2) and self-reports (Samples 3–5). Finally, we explored longitudinal changes in network structure in the aftermath of trauma, assessing whether these changes differed by age using the two longitudinal samples (Samples 6 and 7).

Although symptom-level network structures have varied in previous studies, cluster-level patterns appear more consistent. Our hypothesis therefore, based on the consistent structure of the DSM, was for stability of self-report networks from elementary-school age through to adulthood, with greater centrality and connectivity for reexperiencing symptoms. Given the poor concordance between child- and parent-reports, we expected differences between caregiver- and self-report networks in children and adolescents, hypothesising that self-reports would again reflect greater centrality and connectivity of reexperiencng symptoms, whereas caregiver reports would assign a more integral role to the more observable hyperarousal symptoms. We had no clear hypotheses about differences between preschool- and school-aged children, based on caregiver reports.

## Results

Full networks for Samples 1–5 are shown in Figure 1. Node size reflects symptom centrality, with more interconnected symptoms represented by larger nodes. Edge width indicates connection strength, estimated as regularised partial correlations using the Graphical Least Absolute Shrinkage and Selection Operator (GLASSO). Rings surrounding each node indicate the proportion of variance explained by all other symptoms in the network. The prevalences of PTSD symptoms across clusters in both cross-sectional and longitudinal samples are presented in Table 2.

**Figure 1.**
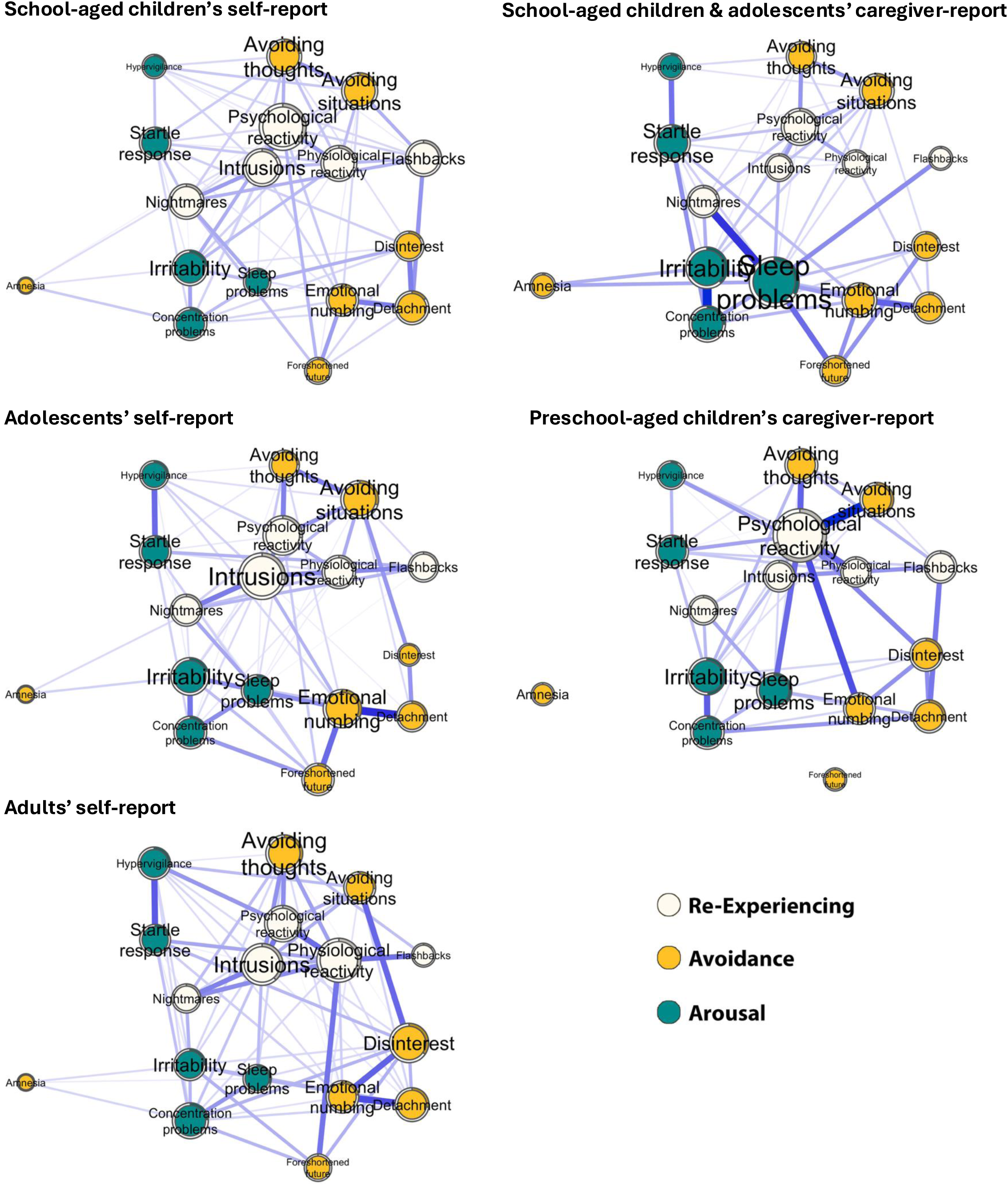
Full symptom networks of samples 1-5, depicting partial correlations between symptoms. Node size indicates centrality in the network. White, orange, and green nodes represent re-experiencing, avoidance, and hyperarousal symptoms, respectively. Blue continuous edges denote significant positive partial correlations, with edge weight (width) reflecting the strength of the partial correlation. Contour circles indicate the predictability of each node based on all other nodes in the network.

**Table 2.**
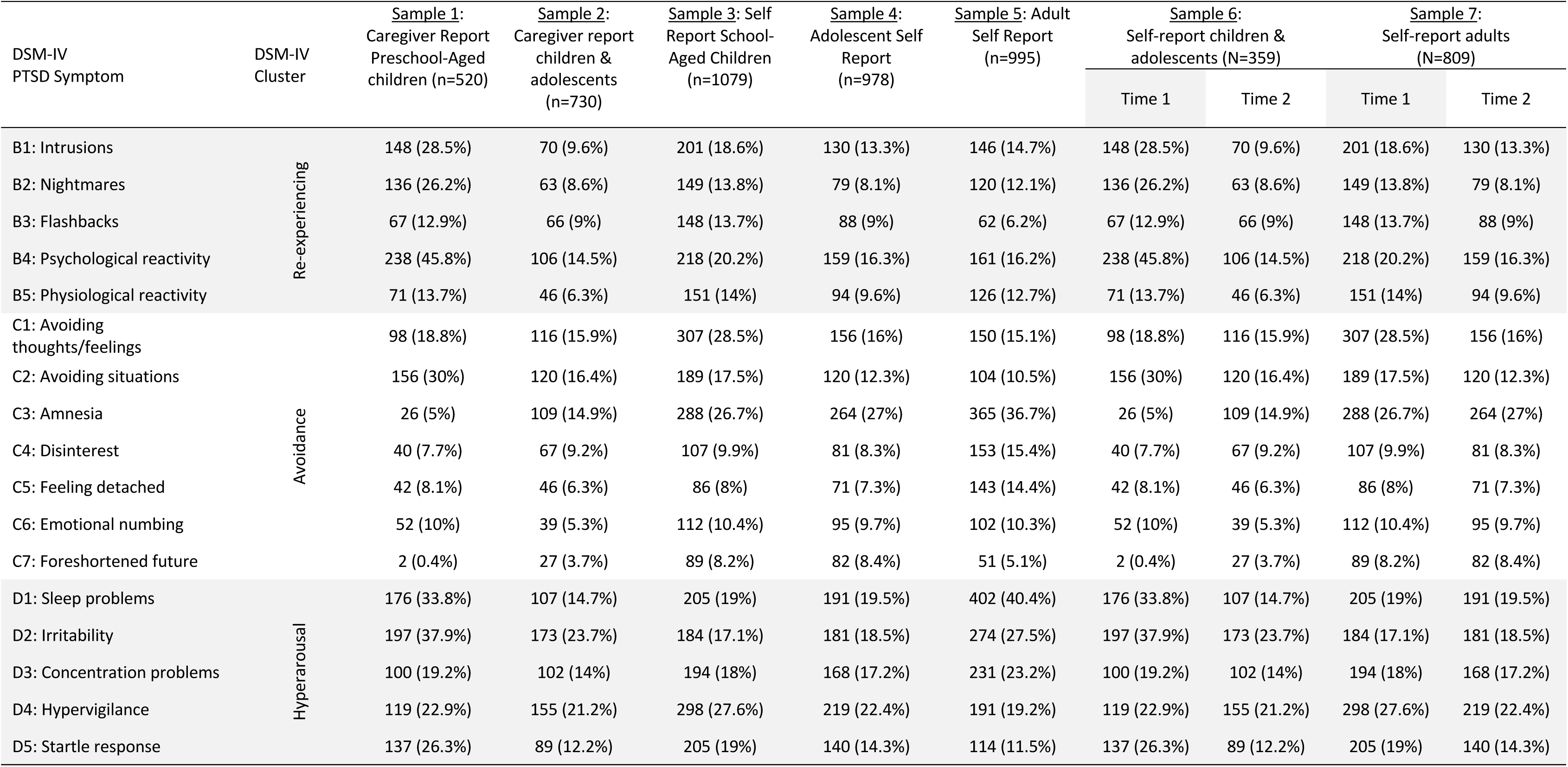
Frequency (n (%)) of PTSD DSM-IV symptom endorsement by sample.

| DSM-IV<br>PTSD Symptom | DSM-IV<br>Cluster | Sample 1:<br>Caregiver Report<br>Preschool-Aged<br>children (n=520) | Sample 2:<br>Caregiver report<br>children &<br>adolescents<br>(n=730) | Sample 3: Self<br>Report School-Aged Children<br>(n=1079) | Sample 4:<br>Adolescent Self<br>Report<br>(n=978) | Sample 5: Adult<br>Self Report<br>(n=995) | Sample 6:<br>Self-report children &<br>adolescents (N=359) |  | Sample 7:<br>Self-report adults<br>(N=809) |  |
| --- | --- | --- | --- | --- | --- | --- | --- | --- | --- | --- |
|  |  |  |  |  |  |  | Time 1 | Time 2 | Time 1 | Time 2 |
| B1: Intrusions | Re-experiencing | 148 (28.5%) | 70 (9.6%) | 201 (18.6%) | 130 (13.3%) | 146 (14.7%) | 148 (28.5%) | 70 (9.6%) | 201 (18.6%) | 130 (13.3%) |
| B2: Nightmares |  | 136 (26.2%) | 63 (8.6%) | 149 (13.8%) | 79 (8.1%) | 120 (12.1%) | 136 (26.2%) | 63 (8.6%) | 149 (13.8%) | 79 (8.1%) |
| B3: Flashbacks |  | 67 (12.9%) | 66 (9%) | 148 (13.7%) | 88 (9%) | 62 (6.2%) | 67 (12.9%) | 66 (9%) | 148 (13.7%) | 88 (9%) |
| B4: Psychological reactivity |  | 238 (45.8%) | 106 (14.5%) | 218 (20.2%) | 159 (16.3%) | 161 (16.2%) | 238 (45.8%) | 106 (14.5%) | 218 (20.2%) | 159 (16.3%) |
| B5: Physiological reactivity |  | 71 (13.7%) | 46 (6.3%) | 151 (14%) | 94 (9.6%) | 126 (12.7%) | 71 (13.7%) | 46 (6.3%) | 151 (14%) | 94 (9.6%) |
| C1: Avoiding<br>thoughts/feelings | Avoidance | 98 (18.8%) | 116 (15.9%) | 307 (28.5%) | 156 (16%) | 150 (15.1%) | 98 (18.8%) | 116 (15.9%) | 307 (28.5%) | 156 (16%) |
| C2: Avoiding situations |  | 156 (30%) | 120 (16.4%) | 189 (17.5%) | 120 (12.3%) | 104 (10.5%) | 156 (30%) | 120 (16.4%) | 189 (17.5%) | 120 (12.3%) |
| C3: Amnesia |  | 26 (5%) | 109 (14.9%) | 288 (26.7%) | 264 (27%) | 365 (36.7%) | 26 (5%) | 109 (14.9%) | 288 (26.7%) | 264 (27%) |
| C4: Disinterest |  | 40 (7.7%) | 67 (9.2%) | 107 (9.9%) | 81 (8.3%) | 153 (15.4%) | 40 (7.7%) | 67 (9.2%) | 107 (9.9%) | 81 (8.3%) |
| C5: Feeling detached |  | 42 (8.1%) | 46 (6.3%) | 86 (8%) | 71 (7.3%) | 143 (14.4%) | 42 (8.1%) | 46 (6.3%) | 86 (8%) | 71 (7.3%) |
| C6: Emotional numbing |  | 52 (10%) | 39 (5.3%) | 112 (10.4%) | 95 (9.7%) | 102 (10.3%) | 52 (10%) | 39 (5.3%) | 112 (10.4%) | 95 (9.7%) |
| C7: Foreshortened future |  | 2 (0.4%) | 27 (3.7%) | 89 (8.2%) | 82 (8.4%) | 51 (5.1%) | 2 (0.4%) | 27 (3.7%) | 89 (8.2%) | 82 (8.4%) |
| D1: Sleep problems | Hyperarousal | 176 (33.8%) | 107 (14.7%) | 205 (19%) | 191 (19.5%) | 402 (40.4%) | 176 (33.8%) | 107 (14.7%) | 205 (19%) | 191 (19.5%) |
| D2: Irritability |  | 197 (37.9%) | 173 (23.7%) | 184 (17.1%) | 181 (18.5%) | 274 (27.5%) | 197 (37.9%) | 173 (23.7%) | 184 (17.1%) | 181 (18.5%) |
| D3: Concentration problems |  | 100 (19.2%) | 102 (14%) | 194 (18%) | 168 (17.2%) | 231 (23.2%) | 100 (19.2%) | 102 (14%) | 194 (18%) | 168 (17.2%) |
| D4: Hypervigilance |  | 119 (22.9%) | 155 (21.2%) | 298 (27.6%) | 219 (22.4%) | 191 (19.2%) | 119 (22.9%) | 155 (21.2%) | 298 (27.6%) | 219 (22.4%) |
| D5: Startle response |  | 137 (26.3%) | 89 (12.2%) | 205 (19%) | 140 (14.3%) | 114 (11.5%) | 137 (26.3%) | 89 (12.2%) | 205 (19%) | 140 (14.3%) |

### Self-Reports: School-aged Children vs Adolescents vs Adults

Aligned with our first hypothesis that self-reports would exhibit a consistent structure from elementary school through adulthood, the three self-report networks were highly similar (Figure 1, left panels). In all three, intrusions (weight [W]=0.90±0.09) and psychological reactivity (W=0.87±0.09), both from the re-experiencing cluster, emerged as the strongest nodes, whereas amnesia (W=0.17±0.04) from the avoidance cluster was consistently the weakest node.

To examine cluster centrality, we conducted a mixed-effects Cluster (within-group; reexperiencing, avoidance, hyperarousal) by Age Group (between-groups; school-aged children, adolescents, adults) ANOVA, adjusted for false discoveries^45^. There were no significant differences in cluster centrality within or between the age groups (Cluster: F_(2,14)_=1.06, η_p_^2^=0.13, p_adj_=.56); Age Group: F_(2,28)_=1.29, η_p_^2^=0.08, p_adj_=.56; Age Group x Cluster: F_(4,28)_=0.44, η_p_^2^=0.06, p_adj_=.78).

We next examined the overall connectivity of the networks using a mixed-effects Cluster by Age Group by Connectivity Type (within-group; within-cluster connectivity, between-cluster connectivity) ANOVA. There was no main effect of Age Group (F_(2,390)_=0.16, η^2^_p_ =0.00, p_adj_=.98). There was a significant main effect of Connectivity Type (F_(1,390)_=38.1, η^2^_p_=0.09, p_adj_<.0001), and for specific cluster combinations (e.g., within re-experiencing, within arousal, re-experiencing with avoidance, etc; F_(4,390)_=4.46, η^2^_p_=0.04, p_adj_=.004), but no interaction effects involving Age Group (Age Group x Connectivity Type: F_(2,390)_=1.09, η^2^_p_=0.01, p_adj_=.47; Age Group x Cluster Combination: F_(8,390)_=0.23, η^2^_p_=0.00, p_adj_=.99; Figure 2). Deconstructing the main effect of Connectivity Type showed a consistent pattern across samples: within-cluster connectivity was significantly stronger than between-cluster connectivity (B=2.41, 95% CI=1.72–3.10, p_adj_<.0001).

**Figure 2.**
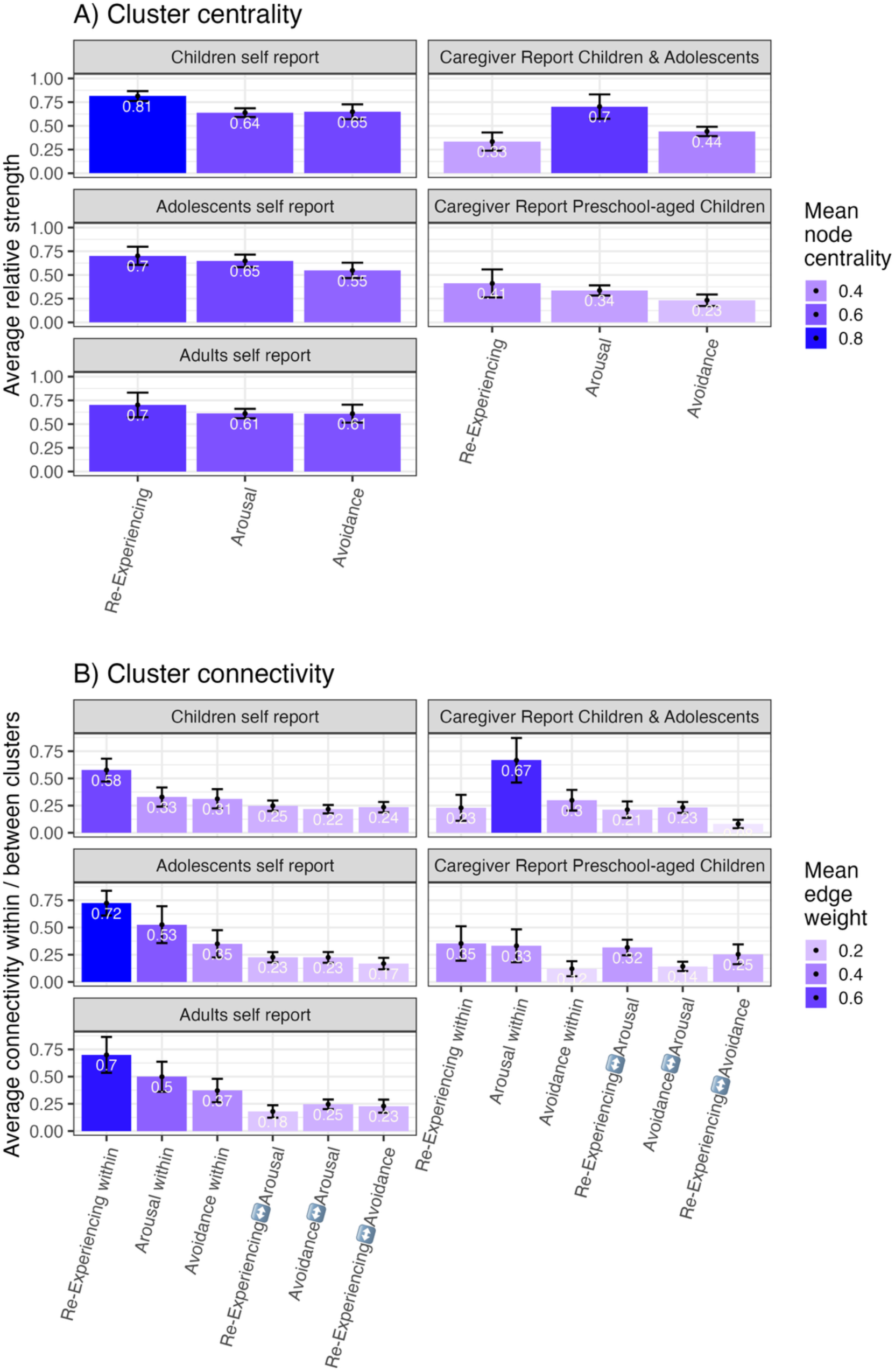
Cluster centrality and cluster connectivity across Samples 1-5. Numbers indicate mean centrality/connectivity value. Error bars represent standard errors. Colors correspond to centrality/connectivity values.

As hypothesized, increased connectivity was most pronounced for the re-experiencing cluster, which showed the highest within-cluster connectivity across all age groups and exceeded every other connectivity pattern (B=5.63, 95% CI=3.69–7.58, all p_adj_ < .0001). Arousal within-cluster connectivity was also significantly stronger than other patterns, except for within re-experiencing (B=2.41, 95% CI=0.86–3.96, all p_adj_=.021). Figures 3 and 4 present the post-hoc comparisons of cluster centrality and connectivity within and between the different age groups in the self-report data.

**Figure 3.**
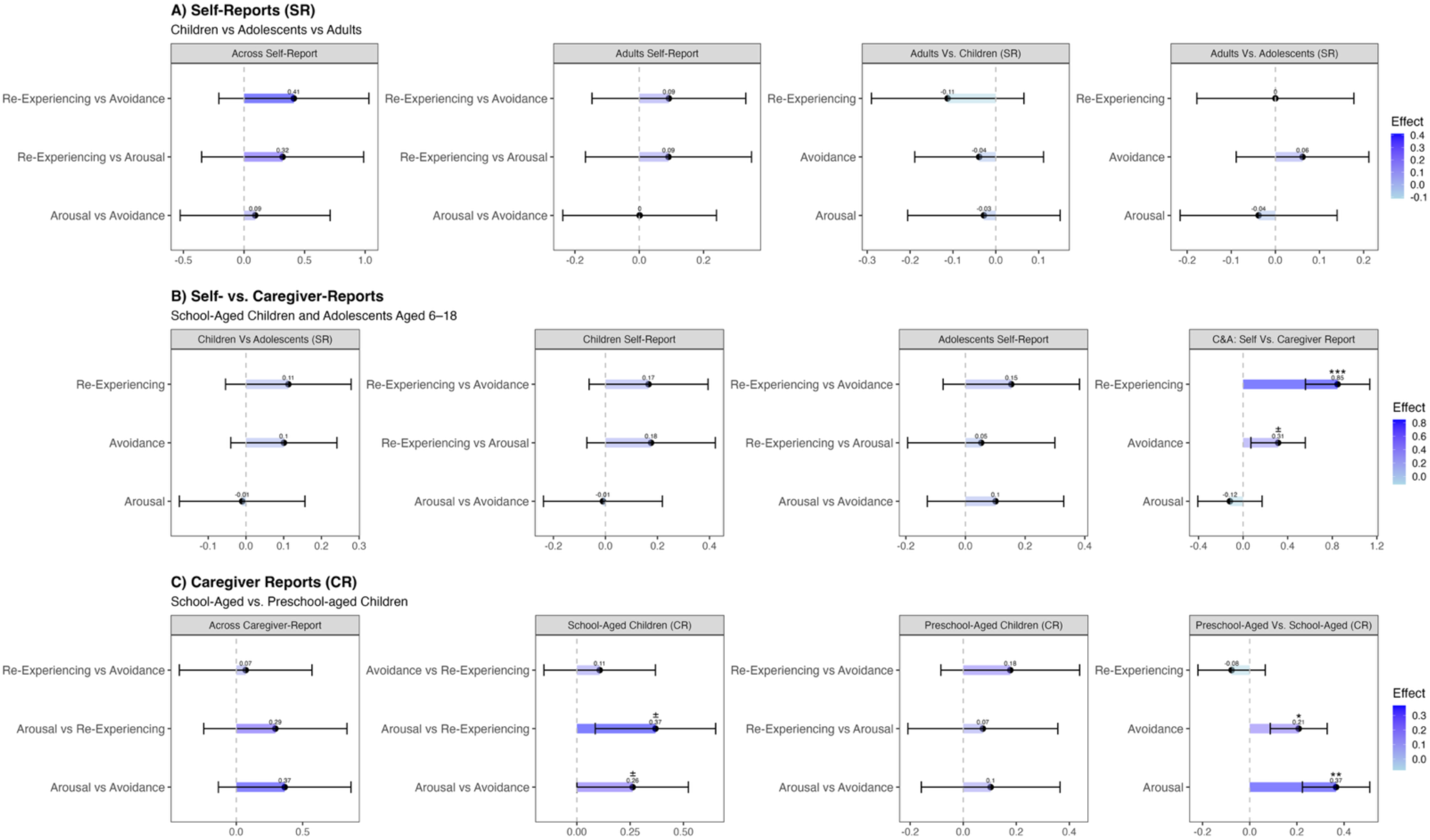
Centrality contrasts in Samples 1-5. Numbers represent effect size (unstandardized B estimate). Error bars represent 95% confidence interval (before adjusting for multiple comparisons). Colours correspond to effect size. * Adjusted p < .05; ** adjusted p < .01; *** adjusted p < .001; ^±^ unadjusted p < .05

**Figure 4.**
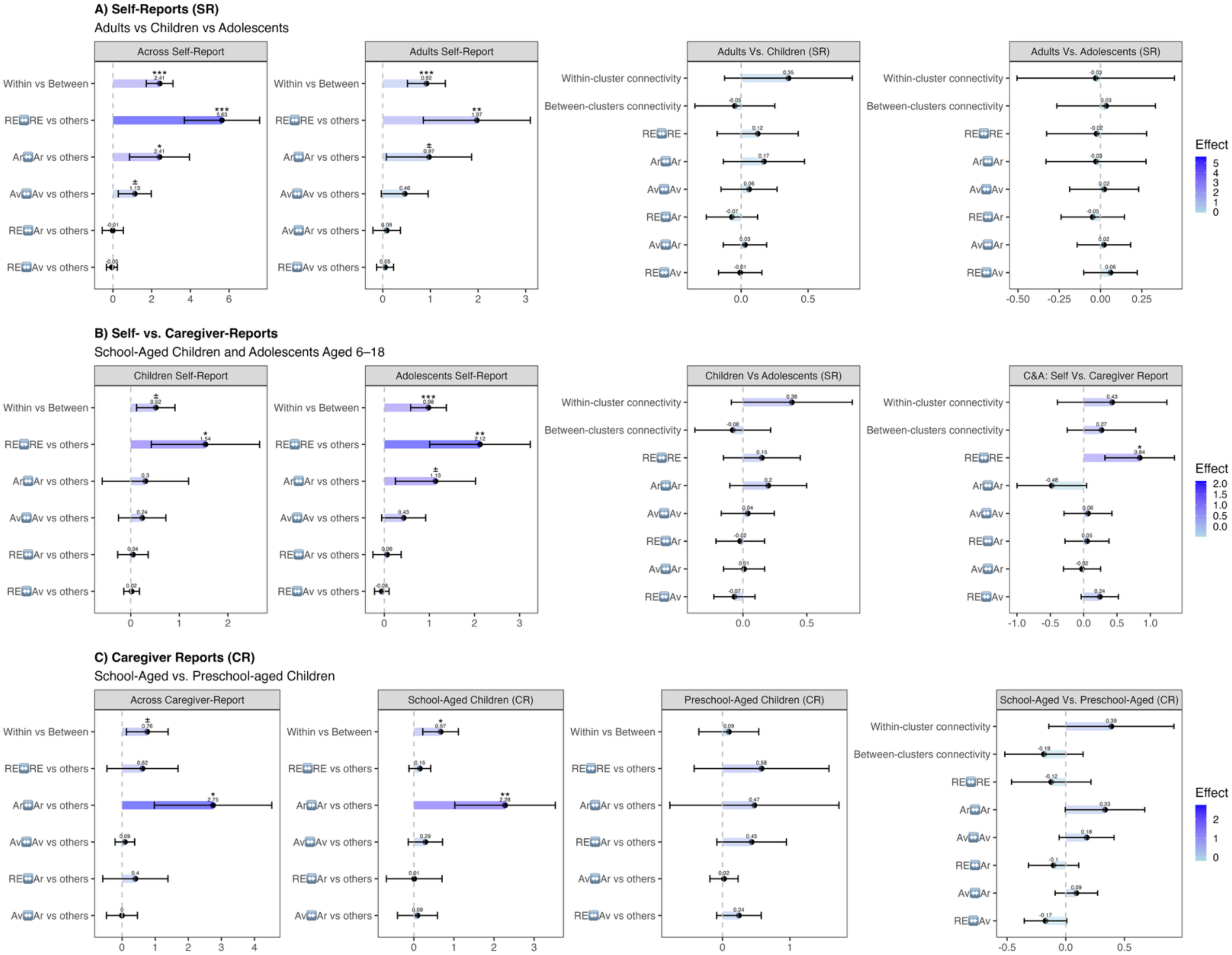
Connectivity contrasts in samples 1-5. Numbers represent effect size (unstandardized B estimate). Error bars represent 95% confidence interval (before adjusting for multiple comparisons). Colours correspond to effect size. * adjusted p < .05; ** adjusted p < .01; *** adjusted p < .001; ^±^ unadjusted p < .05. Abbreviations. RE – Re-experiencing, Ar – Arousal, Av – Avoidance.

### Self- vs. Caregiver-Reports (for School-Aged-Children and Adolescents Aged 6–18)

Aligned with our second hypothesis, school-aged-children’s and adolescents’ symptom networks differed noticeably depending on whether symptoms were caregiver-reported or self-reported (Figure 1). While self-reported networks showed the re-experiencing symptom cluster as most central (specifically, intrusions and psychological reactivity), caregiver reports emphasised the arousal symptom cluster, with sleep problems (W=1.00) and irritability (W=0.89) showing the highest strength centrality.

A mixed-effects ANOVA comparing the three samples (school-aged-children and adolescents: caregiver report, school-aged-children: self-report, and adolescents: self-report) showed a significant effect of Reporter Identity on symptom cluster’s centrality (F_(1,31)_=19.62, η²_p_=0.39, p_adj_=.0003) and a Cluster-by-Reporter interaction (F_(2,31)_=11.4, η²_p_=0.42, p_adj_=.0003). In post-hoc comparisons, re-experiencing symptoms were more central in self-reports than caregiver-reports (B=0.85, 95% CI=0.56–1.14, p_adj_<.0001). Differences in avoidance centrality were non-significant after adjusting for false discoveries (p=.013, p_adj_=.097), and no differences emerged for arousal symptoms’ centrality (p_adj_=.46; Figure 3, upper panel).

Within caregiver reports, contrasting arousal symptoms’ centrality (i.e., the most central symptom cluster among caregivers) against other symptom clusters showed significantly stronger centrality for arousal (B=0.31, 95% CI=0.07,0.54, p=0.014). Similarly, within children’s and adolescents’ self-reports, contrasting re-experiencing symptoms (most central in this group) against other symptoms revealed significantly greater strength (B=0.14, 95% CI=0.01,0.27, p=0.039). No correction for multiple comparisons was applied, as only a single contrast was performed in each case.

Regarding cluster connectivity (Figure 2, lower panel), significant effects were found for Connectivity Type (i.e., within-cluster vs between-clusters; F_(1,396)_=32.3, η²_p_=0.08, p_adj_<.0001), Cluster Pair (i.e., the specific clusters connected; F_(4,396)_=2.94, η_p_^2^=0.03, p_adj_=.034), and a Reporter-by-Cluster Pair interaction (F_(4,396)_=3.62, η_p_^2^=0.04, p_adj_=.016).

In post-hoc comparisons (Figure 4, central panels) within each sample, within-cluster-connectivity within re-experiencing symptoms was significantly stronger than all other connectivity patterns in school-aged-children’s (B=1.54, 95% CI=0.42–2.65, p_adj_=.048) and adolescents’ self-reports (B=2.12, 95% CI=1.01–3.24, p_adj_=.003), whereas in caregivers’ reports, arousal intra-connectivity was significantly stronger than all other patterns (B=2.28, 95% CI=1.02–3.53, p_adj_=.006). Between samples, re-experiencing within-cluster connectivity was stronger in self-reports than in caregiver-reports (B=0.84, 95% CI=0.32–1.36, p_adj_=.017).

### Caregiver-Reports: School-Aged vs. Preschool-Aged Children

Significant centrality effects were found for Age Group (F_(1,14)_=22.4, η²ₚ=0.62, p_adj_=.001) and for the Cluster × Age Group interaction (F_(2,14)_=11.4, η²ₚ=0.62, p_adj_=.002). In post-hoc comparisons, arousal (B=0.37, 95% CI=0.22–0.51, p_adj_=.001) and avoidance (B=0.21, 95% CI=0.09–0.33, p_adj_=.029) were significantly more central in networks of caregivers of school-aged-children and adolescents.

Connectivity analyses initially revealed an effect for specific cluster combinations (F_(4,260)_=2.41, η²ₚ=0.04, p=.049), and marginal effects for Connectivity Type (F_(1,260)_=3.86, η²ₚ=0.01, p=.051) as well as a Connectivity Type-by-Age Group interaction (F_(1,260)_=3.87, η²ₚ=0.01, p=.050), which were attenuated after adjusting for multiple comparisons (pₐ_dj_=.084 for all). As previously reported, arousal symptoms exhibited the strongest within-cluster connectivity among caregivers’ report for school-aged-children and adolescents, but this pattern was not evident in caregivers’ report for preschoolers (Figure 4, lower panel).

### Longitudinal Self-Report Networks

Table 2 (rightmost columns) displays symptom frequencies for the longitudinal samples, with corresponding networks shown in Figure 5. The longitudinal analysis examined network changes from T1 (days since trauma: children and adolescents M±SD = 15.95±12.9 days; adults = 5.94±4.71 days) to T2 (children and adolescents = 238.88±75.8 days; adults = 365 days^2^) within each age group (children & adolescents vs. adults) and tested the Time × Age Group interaction to assess whether network evolution differed by Age Group^3^.

**Figure 5.**
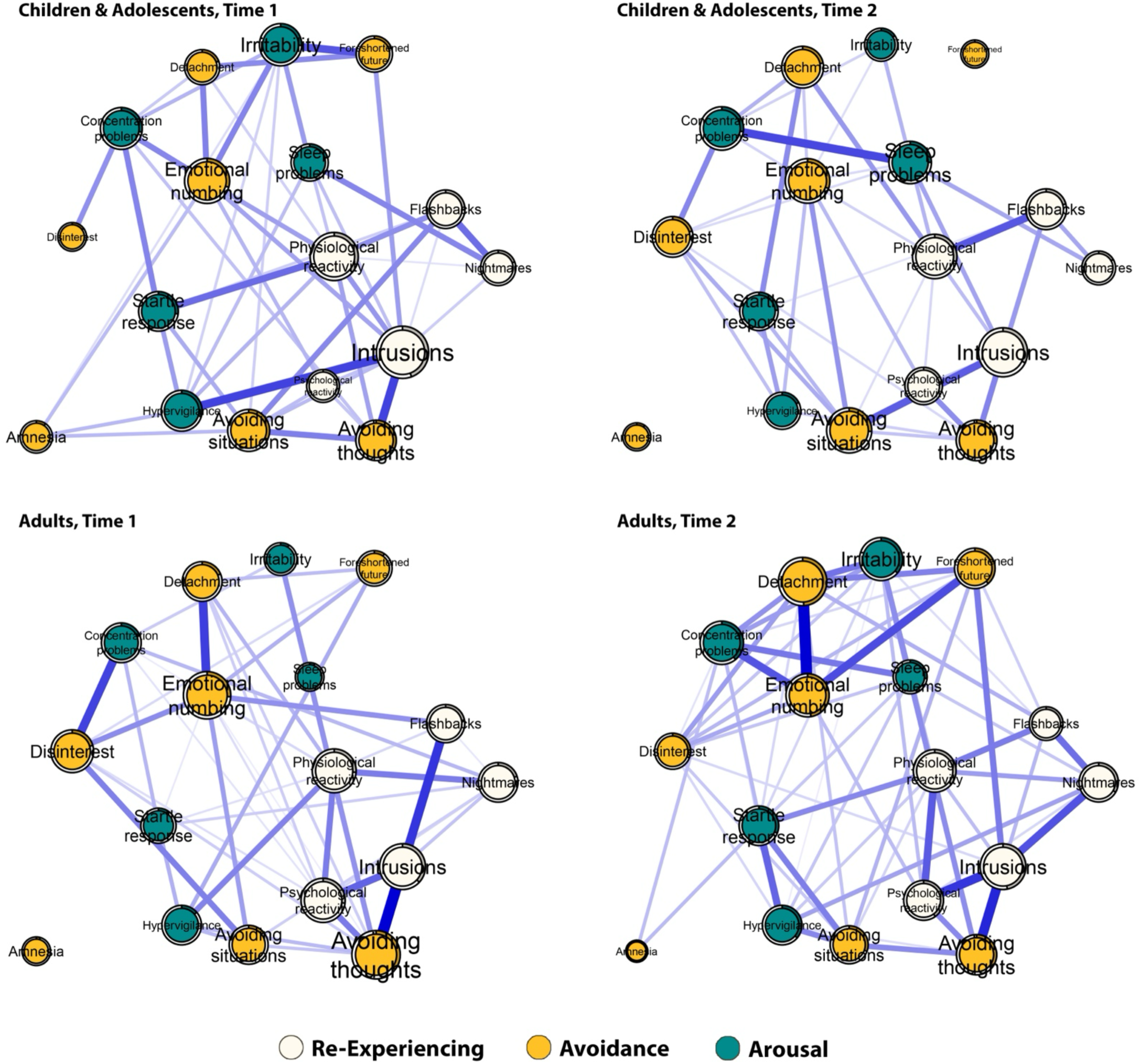
Longitudinal networks for Samples 6-7. Networks depicting partial correlations between symptoms in samples 6-7. Node size represents its centrality in the network. White, orange, and green nodes represent re-experiencing, avoidance, and hyper-arousal symptoms, respectively. Blue continuous edges denote significant and positive partial correlations between symptoms, with edge weight reflecting the strength of partial correlation. Contour circles indicate the predictability of each node by all other nodes in the networks, representing the proportion of variance explained by the remaining network elements.

A mixed-effects ANOVA with Age Group, Time, and Cluster as the independent variables revealed a significant main effect of Age Group on cluster centrality (F_(1,42)_=11.9, η_p_^2^=0.22, p_adj_=.005) and a significant Time by Age Group interaction (F_(1,42)_=31.9, η_p2_=0.43, p_adj_<.0001). Specifically, children and adolescents showed an overall decrease in symptom centrality over time (B=-0.52, 95% CI=-0.83 to -0.19, p_adj_=.006), driven largely by reduced arousal centrality. In contrast, adults showed an overall increase in centrality, particularly for arousal (B=0.45, 95% CI=0.25–0.64, p_adj_=.0001) and avoidance (B=0.18, 95% CI=0.01–0.35, p_adj_=.049) symptoms (Figure 6, upper panels).

**Figure 6.**
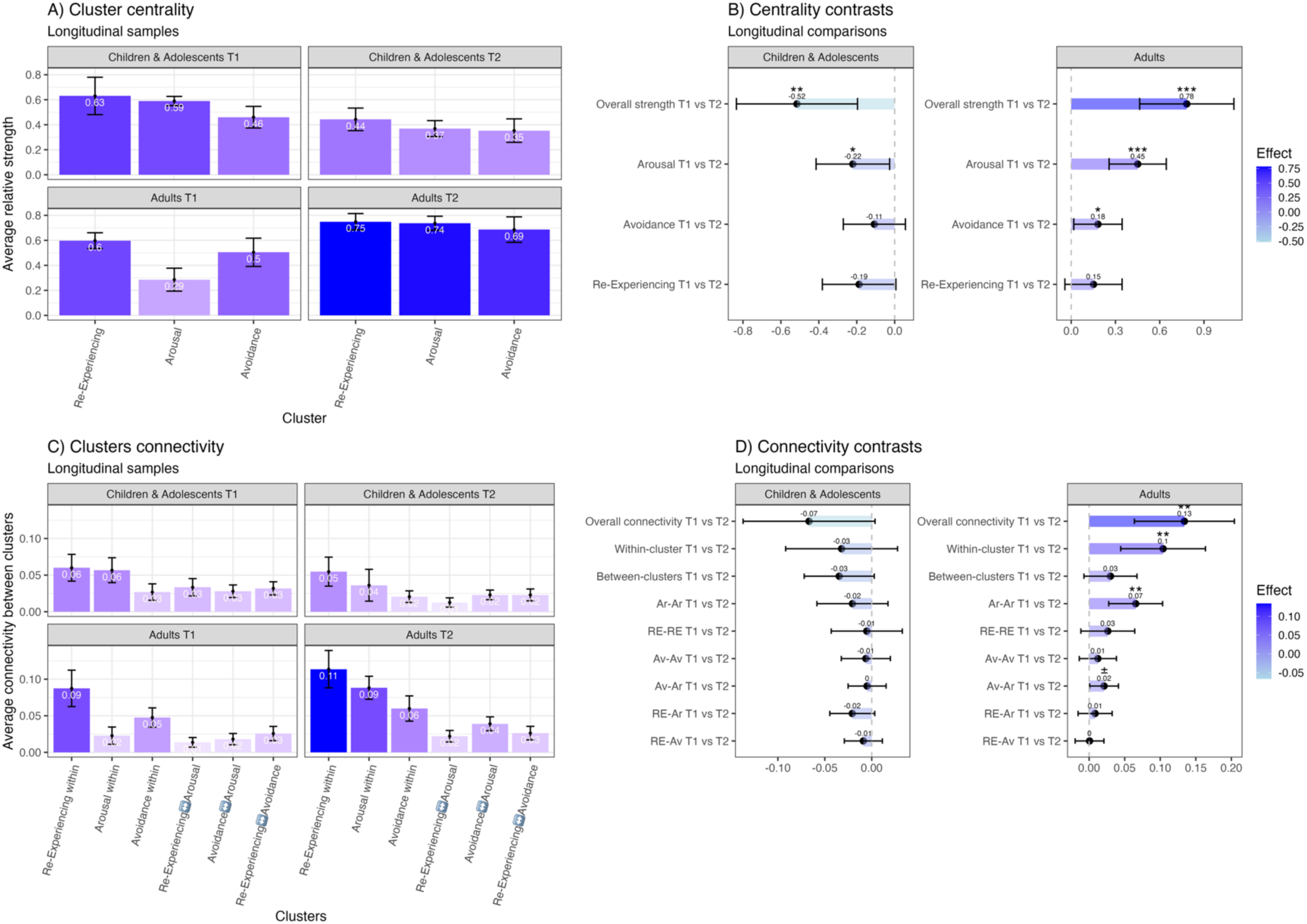
Longitudinal networks – cluster centrality and connectivity. <u>Panels A+C</u>: Numbers indicate mean centrality/connectivity value. Error bars represent standard errors. Colors correspond to centrality/connectivity values. <u>Panels B+D</u>: Numbers represent effect size (unstandardized B estimate). Error bars represent 95% confidence interval (before adjusting for multiple comparisons). Colours correspond to mean value / effect size. * adjusted p < .05; ** adjusted p < .01; *** adjusted p < .001; ^±^ unadjusted p < .05. Abbreviations. RE – Re-experiencing, Ar – Arousal, Av – Avoidance.

Connectivity analyses included a mixed-effects ANOVA with Age Group, Time and Cluster Combination as independent variables, adjusting for specific symptom pairs as random factors^4^ (Figure 6, lower panels). The ANOVA showed a significant Time-by-Age Group interaction (F_(1,390)_=12.4, η_p_^2^=0.03, p_adj_=.002). In post-hoc analyses, while school-aged-children and adolescents did not show significant longitudinal changes in cluster connectivity patterns, adults exhibited a significant overall connectivity increase (B=0.13, 95% CI=0.06–0.20, p_adj_=.004), driven by an increase in within-cluster connectivity (B=0.10, 95% CI=0.04–0.16, p_adj_=.004), and specifically, arousal symptoms’ within-cluster connectivity (B=0.07, 95% CI=0.03–0.10, p_adj_=.004).

### Network Stability

All networks met the recommended stability threshold (correlation-stability coefficient ≥0.25; Supplementary Table S1), with the exception of the longitudinal child and adolescent sample (sample 6) at Time 2. Estimates for Sample 6 at Time 1 as well as caregiver-reported PTSS for children and adolescents (Sample 2) were at the .25 threshold, suggesting caution in interpreting their centrality patterns^46^. See Supplementary Materials (Sup. Figures S1-S18) for visualisations of network stability and edge-weight accuracy.

## Discussion

The current study is the first to concurrently investigate self- and caregiver-reported PTSD symptom networks across preschool-aged children, school-aged children, adolescents, and adults. Unlike the majority of network studies that focused on individual symptoms, our analyses examined higher-level patterns of intra- and inter-connectivity among symptom clusters, capturing broader aspects of PTSD presentation. This approach allowed us to generalize connection patterns across age groups, reducing the risk of overfitting to specific symptoms within particular samples and highlighting recurring patterns.

### Structural similarities and re-experiencing dominance in self-report across age groups

The most striking finding was the consistent strength of re-experiencing connections in self-reports, especially with other re-experiencing symptoms. Among school-aged children, adolescents, and adults, re-experiencing formed the most densely interconnected cluster. Re-experiencing symptoms, such as intrusive memories, nightmares, flashbacks, and psychological reactivity, are inherently internalised and often invisible to external observers. Yet, they are arguably the core of PTSD, a disorder frequently described as a disruption in memory processing^47–50^. This pattern aligns with theoretical models that conceptualise PTSD primarily as a memory-based disorder, where the failure to integrate traumatic experiences into autobiographical memory and a stable sense of self evoke intense affective and physiological responses and perpetuates the post-traumatic response^48^. The clear consistency in cluster centrality and connectivity patterns found across age groups in self-reports suggests that the dynamics of PTSD symptoms may be less age-dependent than commonly assumed, despite some variation in which specific symptoms are most central. These findings highlight the key role of re-experiencing in the persistence of PTSD and point toward prioritising interventions that directly target memory and re-experiencing processes for all post-preschool age groups, including low intensity therapies such as Written Exposure Therapy^51^, or other memory-based interventions. Indeed, recent meta-analyses have identified the most effective interventions for Type-I PTSD (i.e., single event) as those that do exactly this: Trauma-Focused Cognitive Behavioural Therapy (TF-CBT) and Eye Movement Desensitization and Reprocessing (EMDR) for children^52^ and TF-CBT, EMDR, and Prolonged Exposure (PE) for adults^52,53^. These therapies emphasise processing the trauma memory and its sequelae, directly addressing re-experiencing and intrusive symptoms.

### Discrepancy between self- and caregiver reported PTSS in school-aged children and adolescents

In contrast, caregiver reports for children and adolescents showed arousal symptoms as the most strongly interconnected, primarily within their own cluster and less so with other clusters.

These divergent patterns bring to question the reporter’s degree of access, or lack thereof, to internal states, and suggest that externally observable behaviours driven by hyper-arousal are most salient and often most disruptive to parents’ daily functioning. Together, this disparity provides insight into the well-documented discrepancy between caregiver and self-reports in child and adolescent psychiatry^35,37–40^.

Arousal symptoms, including sleep problems, irritability, poor concentration, and exaggerated startle response, are easier to observe and link together than re-experiencing or avoidance symptoms and tend to be what caregivers and others in the individual’s environment (e.g., teachers) most readily notice. Re-experiencing and avoidance symptoms are typically expressed more subtly or internally, and caregivers may not consistently recognise them as trauma-related. It is therefore unsurprising that in caregiver reports, arousal emerged as the most influential cluster, displaying the greatest within-cluster covariance. Re-experiencing symptoms, in contrast, appear less coherently reported by caregivers, while in self-report, they consistently emerge as a central and cohesive network. A recent meta-analysis spanning 30 years of PTSD research revealed striking discrepancies in diagnosis rates depending on the informant: 21.9% when based on children’s self-reports versus 9.9% when based on caregiver reports^54^. Although these differences did not reach statistical significance, they highlight the urgent need to better understand child–caregiver reporting discrepancies. Such gaps may critically influence both diagnostic conclusions and treatment strategies. This is particularly critical in public service settings where access to care is often diagnosis-dependent and relies on caregivers’ report. Our results suggest that self-report might be more reliable when it comes to re-experiencing symptoms, but caregivers might be more consistent in reporting arousal symptoms. Of note, we used both self-report questionnaires and diagnostic interviews, which can yield different outcomes. Diagnostic interviews typically provide more accurate PTSD diagnoses by allowing detailed symptom evaluation, and may help caregivers to more accurately identify re-experiencing symptoms, especially in young children where trauma-related behaviours are easily misinterpreted^55^.

### Differences in caregiver-report for toddlers and preschoolers versus school-aged children

Unlike the consistent self-report patterns observed across age groups, caregiver-reports revealed distinct differences between preschoolers and older school-aged-children and adolescents. Caregiver-reported PTSS for preschool-aged children aged 2 to 6 years did not show reliable differences in intra- or inter-cluster connectivity for arousal, re-experiencing, or avoidance symptoms, despite avoidance symptoms descriptively appearing less connected within the cluster. This may suggest that in early childhood PTSD symptoms do not yet form distinct DSM-like clusters, consistent with the lower symptom threshold for avoidance in the DSM-5 for preschool children. In contrast, caregiver-reports for school-aged children and adolescents showed significantly stronger intra-connectivity within the arousal cluster than any other pattern, suggesting an age-related shift in symptom cluster presentation as perceived by caregivers. Despite a distinct symptom profile underpinning different diagnostic criteria for young children, research examining PTSD network structures in preschool-aged children is notably scarce, making our findings especially important for extending the evidence base in this understudied age group. Bartels et al.^56^ highlighted the centrality of fear and anger, and their association with clinginess, in a large sample of 541 trauma-exposed children aged 1 to 6 years. Cervin et al.^57^ identified a central dyad of intrusions and avoidance symptoms among 75 trauma-exposed children aged 3 to 7 years, with externalizing behaviours related to caregiver stress. To the best of our knowledge, this is the first study to clarify the differences in PTSD network structure between preschoolers and older age groups.

### Differential longitudinal structural shifts in children and adults’ networks

Our longitudinal analyses of self-reported data revealed opposite trends between adults and younger populations. As time passed since the traumatic event, adult symptom networks grew denser, whereas those of children and adolescents became sparser, most notably in relation to arousal symptoms. Among adults, the centrality of arousal increased over time, showing the largest effect and echoing Segal et al^42^. Specifically, arousal symptoms become more intra-connected over time. In contrast to adults, arousal symptoms in children and adolescents decreased in overall centrality over time, without specific changes in their intra- or inter-cluster connectivity. Arousal symptoms are understood as a psycho-biophysiological response to perceived threat following the traumatic event^58^, and are often ‘stickier’, meaning they may persist longer and are more resistant to natural recovery than other symptoms^59^. Such natural resolution over time is considered essential for trauma recovery, while their consolidation into a chronic state is a hallmark of extended posttraumatic stress^60^. The apparent decline in arousal centrality among children and adolescents may reflect their heightened neuroplasticity, which supports greater flexibility in reorganizing fear circuitry and facilitates more rapid restructuring of traumatic memories and biological threat responses^61^. This neurodevelopmental advantage may contribute to faster symptom attenuation and a reduced degree of influence between arousal symptoms and other PTSD symptoms.

Additionally, children and adolescents often benefit from broader and more integrated support networks, such as family, peers, and teachers, who provide resources essential for buffering the impact of trauma and supporting recovery. Indeed, a supportive environment is a well-established resilience factor^62,63^. Unlike the shifts in arousal connectivity, re-experiencing symptoms showed no apparent structural change over time, remaining the most central and internally connected cluster among adults at both time points. This suggests that re-experiencing may stabilise early in the aftermath of trauma, while the co-variation between arousal and avoidance symptoms continues to evolve. Notably, participants in our sample did not receive any intervention, leaving open the important question of whether early treatment might alter these trajectories and influence PTSD severity, chronicity, or recovery, particularly given evidence that early intervention and supportive environment are crucial for PTSD prevention^64^.

### Limitations and conclusions

The current investigation had several limitations. First, although we hypothesised self-report similarity and caregiver-report discrepancies, elucidation of the specific patterns of symptom-cluster connectivity were exploratory rather than confirmatory as this was the first study to examine these questions. Although we corrected for multiple comparisons and applied stringent significance criteria, future studies should aim to replicate and confirm these results in independent samples to confirm their stability. Nonetheless, the consistent cluster-level patterns observed across mainly independent samples suggest a consistent structure of symptom connectivity across samples and age groups. A second limitation concerns potential differences in symptom severity across samples, as our focus was on symptom presence rather than intensity and we relied on harmonized scores from diverse measures (see Methods section and Sup. Table S3). Previous studies have addressed this by including additional severity measures, which was not feasible in the current study. In our longitudinal analyses, we accounted for symptom severity over time by regressing out time effects from symptom presence (i.e., controlling for overall increases or decreases). However, it remains possible that symptom severity confounded the network structures. Third, some of our data were collected prior to the introduction of DSM-5, preventing inclusion of the “negative alterations in cognition and mood” cluster, which likely influences network structure. Beyond this, the reliance on DSM-defined symptoms without broader cognitive, behavioral, or emotional mechanisms, is a limitation in itself. The DSM is designed for diagnostic consistency, but this categorical approach may be at odds with the conceptualisation of symptoms as components of a dynamic transdiagnostic system^25,65^. Psychological models, such as Ehlers and Clark’s cognitive model of PTSD^10^, may provide a more informative framework for understanding the disorder as an interacting system. Future studies should examine how PTSD manifests across age groups beyond DSM categories, drawing on mechanistic models of the disorder. As our participants originated from the US, UK, and Australia, our findings warrant replication in culturally diverse samples. Likewise, future studies should examine how network structures may differ following single-event versus repeated-trauma exposures. Finally, as our analyses are based on cross-sectional, between-subject networks, the observed associations may not fully reflect within-person dynamics. This is an important consideration, particularly as network science increasingly shifts toward idiographic methods that track symptom fluctuations within individuals over time. However, this limitation does not undermine the value of our approach: between-subject networks remain important for characterising PTSD symptom profiles and estimating the likelihood of co-occurring symptom patterns in traumatized populations.

This is the first study to concurrently examine intra- and inter-cluster PTSD symptom dynamics across the lifespan. Drawing on seven samples, including both self- and caregiver-reports, we demonstrate age-related differences in PTSD presentation and the influence of reporter identity. Our findings highlight the centrality of self-reported re-experiencing symptoms and how these dynamics differ when symptoms are reported by caregivers. They also illustrate differential longitudinal changes in network structure and density between children and adults, and show that caregiver reports for preschoolers display less coherent symptom clustering compared to school-aged children. Together, these insights can inform both clinical assessment and intervention design, particularly when working with diverse age groups and informants.

## Methods

### Participants and Procedure

Data were collated from multiple studies investigating PTSD symptomatology^11,66–70^. Five different samples were derived from these datasets based on pre-defined age groupings. Two of the samples involved caregiver reports of child and adolescent PTSD symptoms while the remaining three involved self-report. All participants were assessed using standardized measures designed to assess post-traumatic stress disorder (PTSD) symptoms according to the Diagnostic and Statistical Manual for Mental Disorders: Fourth Edition (DSM-IV)^71^. A summary of the sample characteristics for the five datasets is provided in Table 1.

### Caregiver-report samples

Sample one comprised data from 520 caregivers of trauma-exposed children aged between 2 and 6.5 years (M_age_=4.73 years, SD=1.11; Sex_female_=216, 41.5%). Of these very young children, 43 (8.3%) met criteria for DSM-IV PTSD, with 158 (30.4%) meeting developmentally-appropriate criteria for PTSD according to the Alternative Algorithm (AA) – a precursor of the Preschool PTSD diagnosis in the DSM-5 developed for this age group^72,73^ (see Supplementary Table S2 for an overview of the algorithm and its relationship to the DSM-5 PTSD preschool subtype), based on caregiver-report.

Sample two comprised 730 caregivers of older trauma-exposed school-aged children and adolescents aged between 6.5 and 18 years of age (M_age_=10.12; SD=2.57; Sex_female_=260, 35.6%). Of this sample, 28 (3.8%) met criteria for DSM-IV PTSD using the appropriate adult-based algorithm.

### Self-report samples

Sample three overlapped with Sample Two and comprised self-reported symptoms of post-traumatic stress in 1079 elementary-school aged children aged between 6.5 and 12 years (M_age=_10.10, SD=1.51; Sex_female_=449, 41.6%). Of this sample, 103 (9.5%) met standard criteria for DSM-IV PTSD.

Sample four also overlapped with Sample two and comprised 978 adolescents aged between 13 and 18 years (M_age_=14.84, SD=1.39; Sex_female_=349, 35.7%) with self-reported posttraumatic stress symptoms. Of these, 74 (7.6%) met criteria for DSM-IV PTSD.

Finally, Sample five was an adult sample, comprising 995 individuals aged 18 and older (M_age_=37.84, SD=13.76; Sex_female=_261; 26.2%), who self-reported their symptoms of post-traumatic stress. Of these, 103 (10.4%) met criteria for a PTSD diagnosis.

Details of the inclusion criteria and recruitment flow for each of the different samples is summarised in the Supplementary Materials (Sup. Figures S19-S21). Briefly, the majority of the Sample one’s preschool-aged child dataset was obtained from these four studies^66,67,69,70^ in conjunction with the Prospective Studies of Acute Child Trauma and Recovery Data Archive (PACT/R)^68^. PACT/R is an international collaborative effort to share and preserve child trauma data. Information about the studies included in this data archive can be found here: https://childtraumadata.org. Samples two and three comprised data acquired from PACT/R in addition to participants in the previously aforementioned studies whose age was above 6.5 years. Sample four was mainly comprised of data from PACT/R and from Bryant et al.^11^, were a participant to be under 18 years of age. Finally, Sample five was comprised mainly of participants from the Bryant et al.,^11^ study and participants from the PACT/R database who were above 18 years of age.

### Participants for longitudinal network evaluations

Two longitudinal samples (see Table 1) of children and adolescents (Sample Six; *N*=359) and adults (Sample Seven; N=809) were extracted from the three self-report cross-sectional samples to include participants for whom we had repeated measures of post-traumatic stress symptoms across at least two time points. For Time 1 we required that participants be assessed within 28 days of having experienced a trauma. This criterion was utilised given it has been defined within the DSM-IV-TR as the point in time in which acute stress reactions occur^71^. We required a minimum time difference of three months (90 days) between Time 1 and follow-up at Time 2 as it has been observed that after three months allows for ‘natural recovery’ to occur following acute symptoms in children and young people^74,75^.

### Harmonisation of posttraumatic stress measures across samples

The different post-traumatic stress measures used in the source studies for the 5 cross-sectional samples are presented in Table 1. A harmonization guide was used to create a standardised database across these different assessment measures. This guide was created as part of the PACT/R project and involved a panel of international leaders in PTSD research identifying the degree to which each question of each assessment measure assessed the relevant DSM-IV PTSD symptom^68^. To standardize across measures, each item was binarized to indicate whether the person had either met or did not meet the criteria. See Supplementary Materials for details on the harmonization process and the derivation of diagnoses for each measure and sample (Sup. Table S3).

### Overview of analytic approach

#### Cross-sectional regularized partial correlation networks

Cross-sectional networks were estimated for samples one to five using Ising Models^76^. Ising Models estimate weighted, undirected networks among binary items, where the pairwise interactions represent conditionally independent associations between nodes, analogous to partial correlations. Using the *IsingFit* package in R^77^, we applied a regularized estimation procedure combining the Graphical Least Absolute Shrinkage and Selection Operator (GLASSO) with an Extended Bayesian Information Criterion (EBIC). The hyperparameter for regularization was set at 0.25, the default in *IsingFit*^78^. GLASSO shrinks all edge weights toward zero and sets small weights exactly to zero, thereby producing a sparse network. The R package *qgraph*^79^ was used to visualize the resulting structures, where thicker links indicate stronger associations, either positive (blue) or negative (red, dotted), and the absence of a link indicates conditional independence after regularization.

#### Network Inference

We quantified node importance using Strength centrality (Degree Centrality), computed with the *bootnet* package^80^. Strength reflects the summed absolute edge weights connected to a node, thus representing how interconnected it is in the network. Strength centrality was normalized for comparability across networks. Node predictability, the shared variance between each node and all of its neighbours, was computed using the *mgm* package in R^81^.

#### Stability

Network Stability for each of the respective networks was computed using the procedures outlined by Epskamp, Borsboom and Fried^46^. The R package *bootnet* was used to assess the stability of strength centrality and the accuracy of edge weights. In line with these recommendations, *bootnet* performed 1000 bootstraps on progressively smaller, randomly drawn subsets of the sample. Edge-weight accuracy was estimated through bootstrapped confidence intervals, and stability was quantified using the correlation-stability (CS) coefficient. Values of the CS-coefficient above 0.50 are considered desirable, whereas values below 0.25 are regarded as insufficient^46^.

#### Visualisation

To visualize the outputs produced by *qgraph*, the layout of the nodes was supervised using the Fruchtermann-Reingold algorithm^82^. For ease of visual comparison, the layout of the networks was restricted using the *‘Average Layout’* command in *qgraph* so each network had a comparable layout. We visualised edge weights on a similar weight by enforcing a unified maximum value for visualisation, which was the strongest edge across all networks. We used node size to represent centrality, so that visibly bigger nodes are more central than smaller ones.

#### DSM Cluster centrality

To comprehensively investigate association patterns within the DSM-IV symptom clusters (i.e., Re-experiencing, Arousal, and Avoidance), we conducted a series of mixed-effects Analyses of Variance (ANOVA) with strength centrality as the dependent variable. Independent variables included reporter identity (self-report vs. caregiver-report) and age group. Reporter Identity and Age Group were modelled as between-person fixed factors, while Symptom Cluster was modelled as a within-person fixed factor. Specific symptoms were modelled as random effects where it enabled the model to converge. Cluster Centrality was computed by averaging the strength of symptoms within each cluster.

In line with our hypotheses, we constructed three cross-sectional models:

1. A comparison of self-report across age groups: school-aged children, adolescents, and adults, conducted using the three self-report samples (3-5).
2. A comparison of self- vs. caregiver-report in school-aged-children and adolescents, using Samples 2-4 (children and adolescents’ caregiver-report, children’s self-report, and adolescents’ self-report).
3. A comparison of caregiver-report for parents of school-aged-children and adolescents vs. parents of preschool-aged children, using Samples 1 (preschool-aged children’s caregiver-report) and 2 (school-aged-children’s and adolescents’ caregiver-report).

To clarify specific effects, we conducted targeted pairwise comparisons following the main analyses. These included comparisons between clusters (e.g., arousal vs. avoidance) within each age group and reporter identity, as well as comparisons of the same cluster across age groups and reporters (e.g., re-experiencing in caregivers’ reports vs. children’s and adolescents’ self-reports), using the *emmeans* package ^83^. The analyses controlled for false discoveries using Benjamini and Hochberg’s False Discovery Rate (FDR) adjustment^45^. FDR adjustment was applied across all comparisons to minimise the risk of a Type I error.

#### DSM-clusters connectivity – intra- vs. inter-connectivity

To analyse connectivity between symptoms belonging to different clusters (inter-connectivity, or Between-Cluster Connectivity) or within the same cluster (intra-connectivity, or Within-Cluster Connectivity), edges in the network were labelled based on the clusters they connected. For example, the edge between intrusions and nightmares is an intrinsic intra-connectivity edge, linking symptoms within the re-experiencing cluster, while the edge between intrusions and irritability is a between-clusters edge, connecting the re**-**experiencing cluster with the arousal cluster. In these analyses, edge weight served as the dependent variable.

We repeated the same analyses described earlier with a few modifications. Specifically, we included Connectivity Type (within- or between-cluster connectivity) and specific cluster-pair^5^ as the fixed independent variables without modelling specific symptoms as random effects, as doing so led to overfitting due to the cluster-pair variable. We followed up with post-hoc comparisons in which each connectivity pattern was compared against all connectivity patterns of lower strength, and adjusted p-values for multiple comparisons using FDR across all comparisons.

#### Longitudinal analyses

Samples Six and Seven allowed us to examine the consistency of the network structure over time and assess potential structural changes from acute to chronic post-traumatic stress reaction, when sampled more than once over extended periods. This analysis was restricted to participants who were measured at two time points. To adjust for degree of overall symptom change over time (symptom severity), we regressed the group-level effect of time on symptom presence (1 or 0) and used a Mixed Graphical Model (MGM) to estimate the network, rather than the Ising model. The MGM is particularly well-suited for this analysis as it can model both continuous and categorical variables within the same framework, making it ideal for the de-trended variables.

As with the previous analyses, we computed DSM-cluster Centrality and Connectivity at each time point, following the same methodology used for the cross-sectional samples. This allowed us to compare the network structures across time and observe whether any significant structural changes occurred.

#### Missing Data

For the network analysis, participants were only included if there were completed measures of each posttraumatic stress symptom and age-related information was available (see Tables 1 and 2, and Supplementary Figure S21 for exclusion criteria). For a small percentage (less than 5% across samples) where 1-3 symptoms were missing, we imputed values using the sample’s symptom mode.

## Data Availability

The datasets used and/or analyzed during the current study are available from the corresponding author upon reasonable request.
Analysis codes are available at Open Science Framework: https://osf.io/syb9v/overview?view_only=17d033727dbf4308a068e63c7e43164e

## Contributors

All authors contributed to, reviewed, and approved the final manuscript.

Conceptualization: DJ, NH-P, AB, TD

Methodology: DJ, NH-P, AB, TD

Software: DJ, NH-P

Validation: NH-P

Formal analysis: NH-P

Investigation: CH, MB, AdH, RB, AM, ADY, PS, RM-S

Resources: CH, MB, AdH, RB, AM, ADY, PS, RM-S, TD

Data curation: DJ, NH-P

Writing – original draft: DJ, NH-P

Writing – review & editing: CH, MB, AdH, AM, ADY, PS, RM-S, AB, TD

Visualization: NH-P

Supervision: AB, TD

Project administration: DJ, NH-P, TD

## Availability of data and materials

The datasets used and/or analyzed during the current study are available from the corresponding author upon reasonable request.

Analysis codes are available at Open Science Framework: https://osf.io/syb9v/overview?view_only=17d033727dbf4308a068e63c7e43164e

## Ethics statement

Ethical approval was not required for this study as it is based on anonymized pre-existing datasets where participants cannot be identified. The data were treated in accordance with the UK Data Protection Act (2018) and no confidential information was accessed.

## Footnotes

1 As some data were collected previous to the introduction of the DSM-5, negative alterations in mood and cognition were not analysed as a cluster.

2 Distribution data on days since trauma at T2 was unavailable for this sample.

3 Overall symptom severity change was controlled, see methods section.

4 As we were interested in cluster-level dynamics, we controlled for baseline differences in specific symptoms within clusters.

5 RE-RE, Ar-Ar, Av-Av, RE-Ar, RE-Av, or Ar-Av; RE = Re-experiencing; Ar = Arousal; Av = Avoidance

