## Supplementary materials 1 for "The Inter-Connectedness of Post-Traumatic Stress Disorder (PTSD) Symptomatology Across the Lifespan"

| **Table S1**. Correlation-stability coefficients for the Strength centrality index | | | | | | | | |
| --- | --- | --- | --- | --- | --- | --- | --- | --- |
| Cross-sectional samples | Sample 1: Caregiver Report Preschool-Aged Children | Sample 2: Caregiver Report Children & Adolescents | | Sample 3: Self-report School-Aged Children | | Sample 4: Self-report Adolescents | | Sample 5: Self-report Adults |
| Strength CS-coefficient | .68 | .25 | | .51 | | .53 | | .63 |
| Longitudinal samples | Sample 6; Time 1:  Self-Report Children and Adolescents | | Sample 6; Time 2:  Self-Report Children and Adolescents | | Sample 7; Time 1:  Self-Report Adults | | Sample 7; Time 2:  Self-Report Adults | |
| Strength CS-coefficient | .25 | | .23 | | .60 | | .64 | |

Note. *CS=Correlation- Stability*

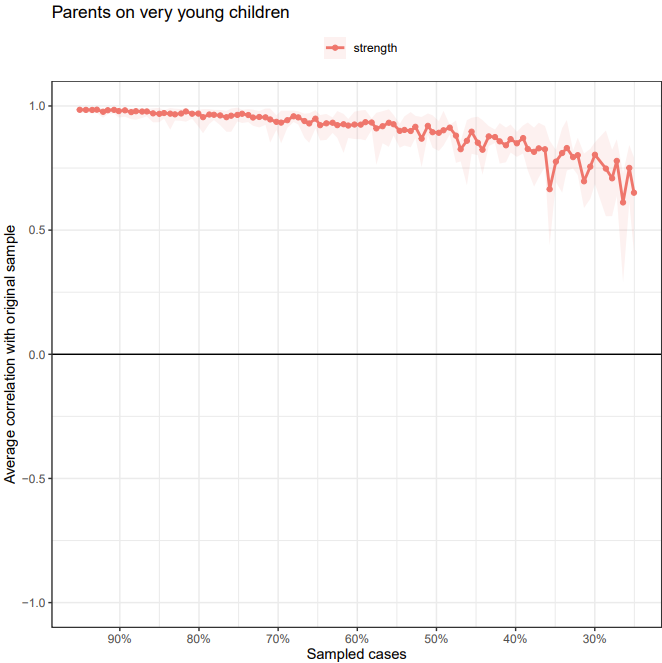

**Figure S1. Subset bootstrap test.** Subsetting bootstrap for the 17 DSM-IV-TR symptom network, estimated based on the caregiver report of preschool-aged children dataset. The table shows the average correlation between centrality indices from the original estimated network.

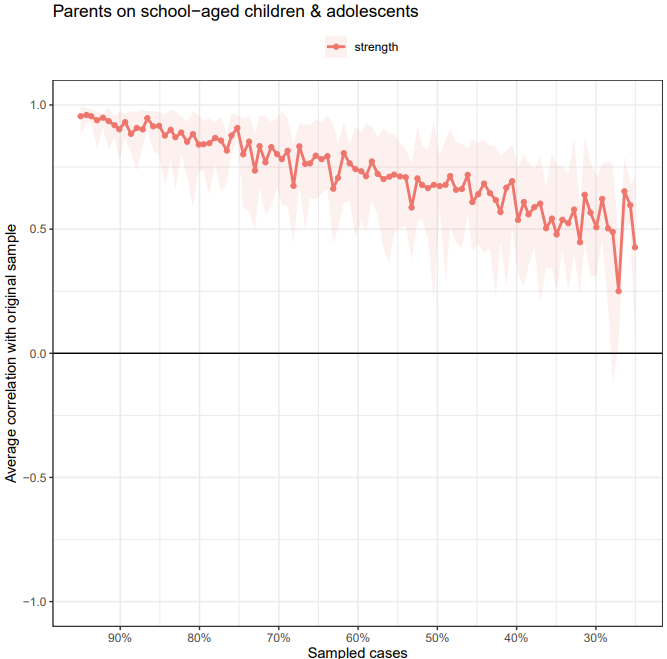

**Figure S2. Subset bootstrap test.** Subsetting bootstrap for the 17 DSM-IV-TR symptom network, estimated based on the caregiver report of children and adolescents dataset. The table shows the average correlation between centrality indices from the original estimated network.

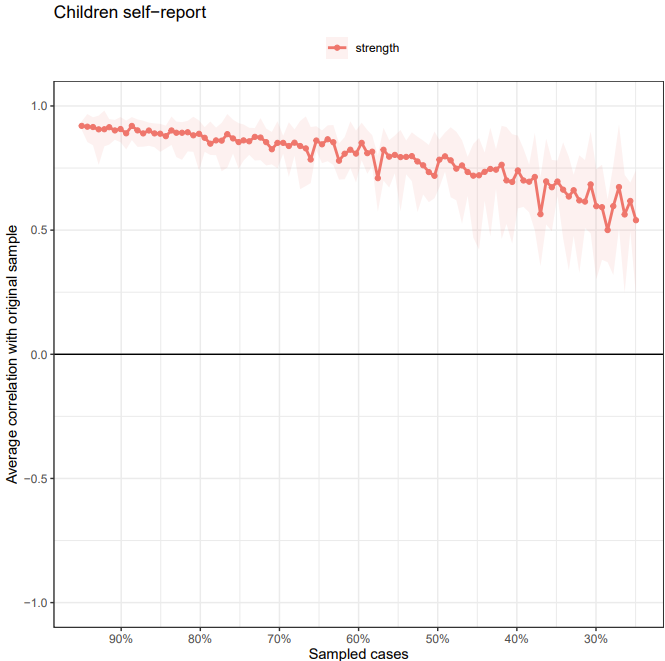

**Figure S3. Subset bootstrap test.** Subsetting bootstrap for the 17 DSM-IV-TR symptom network, estimated based on the self-report of children dataset. The table shows the average correlation between centrality indices from the original estimated network.

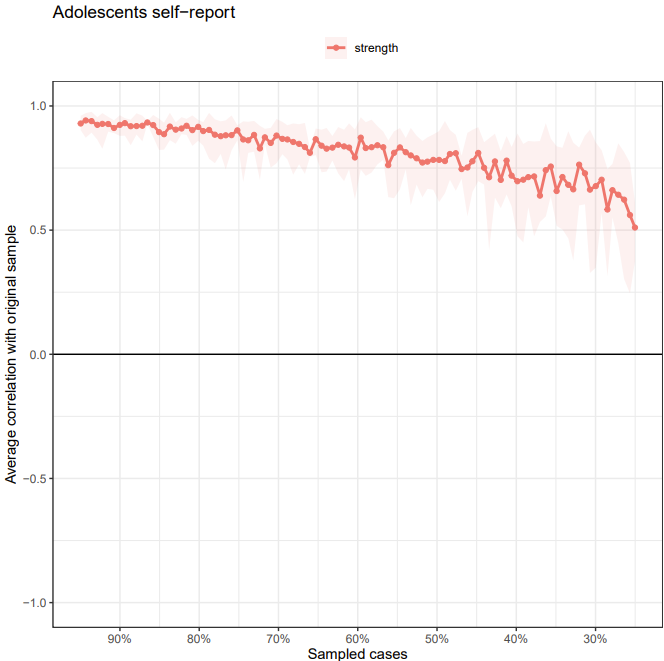

**Figure S4. Subset bootstrap test.** Subletting bootstrap for the 17 DSM-IV-TR symptom network, estimated based on the self-report of adolescents dataset. The table shows the average correlation between centrality indices from the original estimated network.

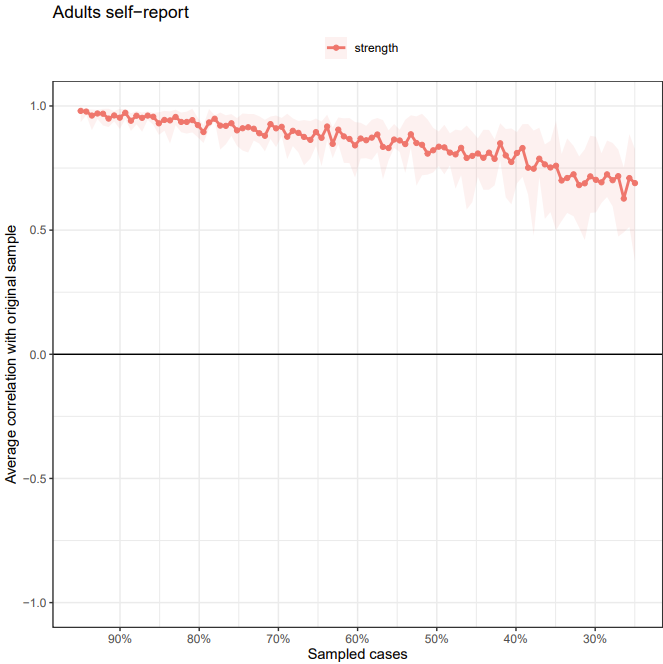

**Figure S5. Subset bootstrap test.** Subsetting bootstrap for the 17 DSM-IV-TR symptom network, estimated based on the self-report of adults dataset. The table shows the average correlation between centrality indices from the original estimated network.

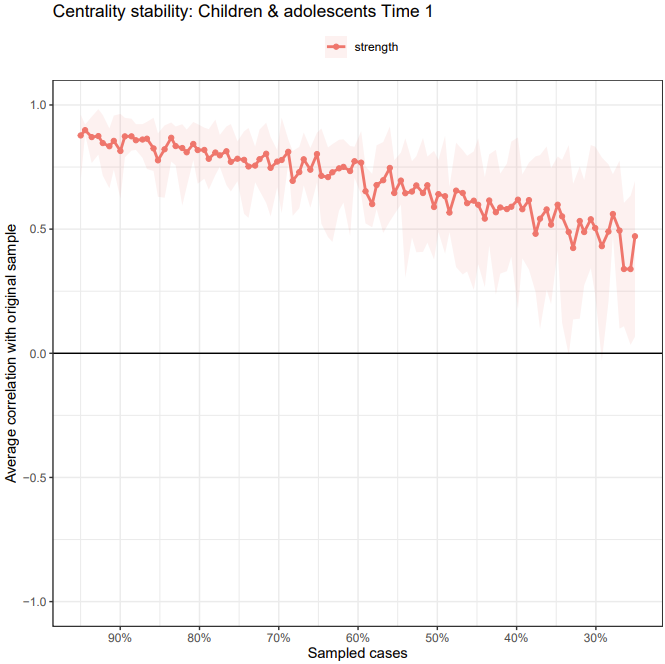

**Figure S6. Subset bootstrap test.** Subsetting bootstrap for the 17 DSM-IV-TR symptom network, estimated based on the self-report of the longitudinal child & adolescent dataset (time 1). The table shows the average correlation between centrality indices from the original estimated network

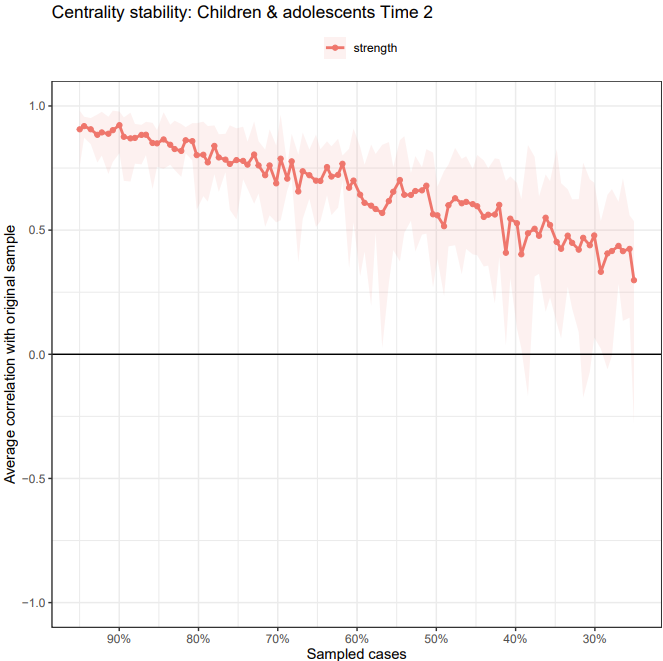

**Figure S7. Subset bootstrap test.** Subsetting bootstrap for the 17 DSM-IV-TR symptom network, estimated based on the self-report of the longitudinal child & adolescent dataset (time 2). The table shows the average correlation between centrality indices from the original estimated network

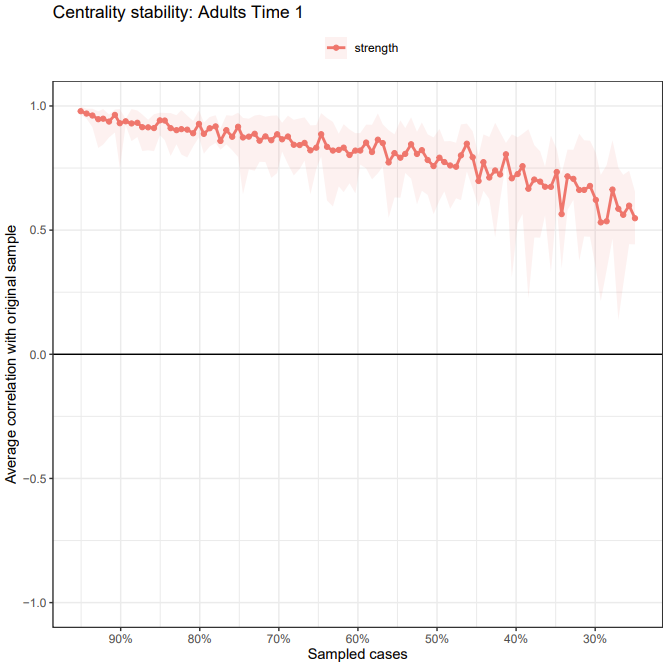

**Figure S8. Subset bootstrap test.** Subsetting bootstrap for the 17 DSM-IV-TR symptom network, estimated based on the self-report of the longitudinal adult dataset (time 1). The table shows the average correlation between centrality indices from the original estimated network.

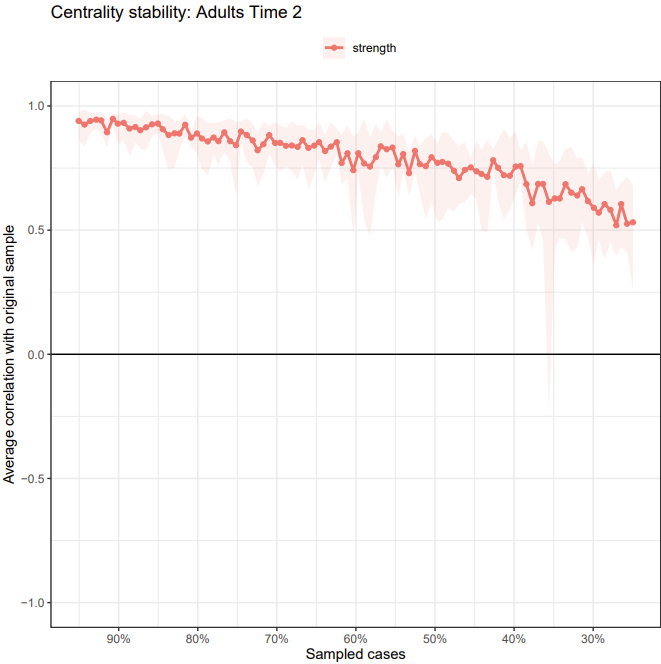

**Figure S9. Subset bootstrap test.** Subsetting bootstrap for the 17 DSM-IV-TR symptom network, estimated based on the self-report of the longitudinal adult dataset (time 2). The table shows the average correlation between centrality indices from the original estimated network.

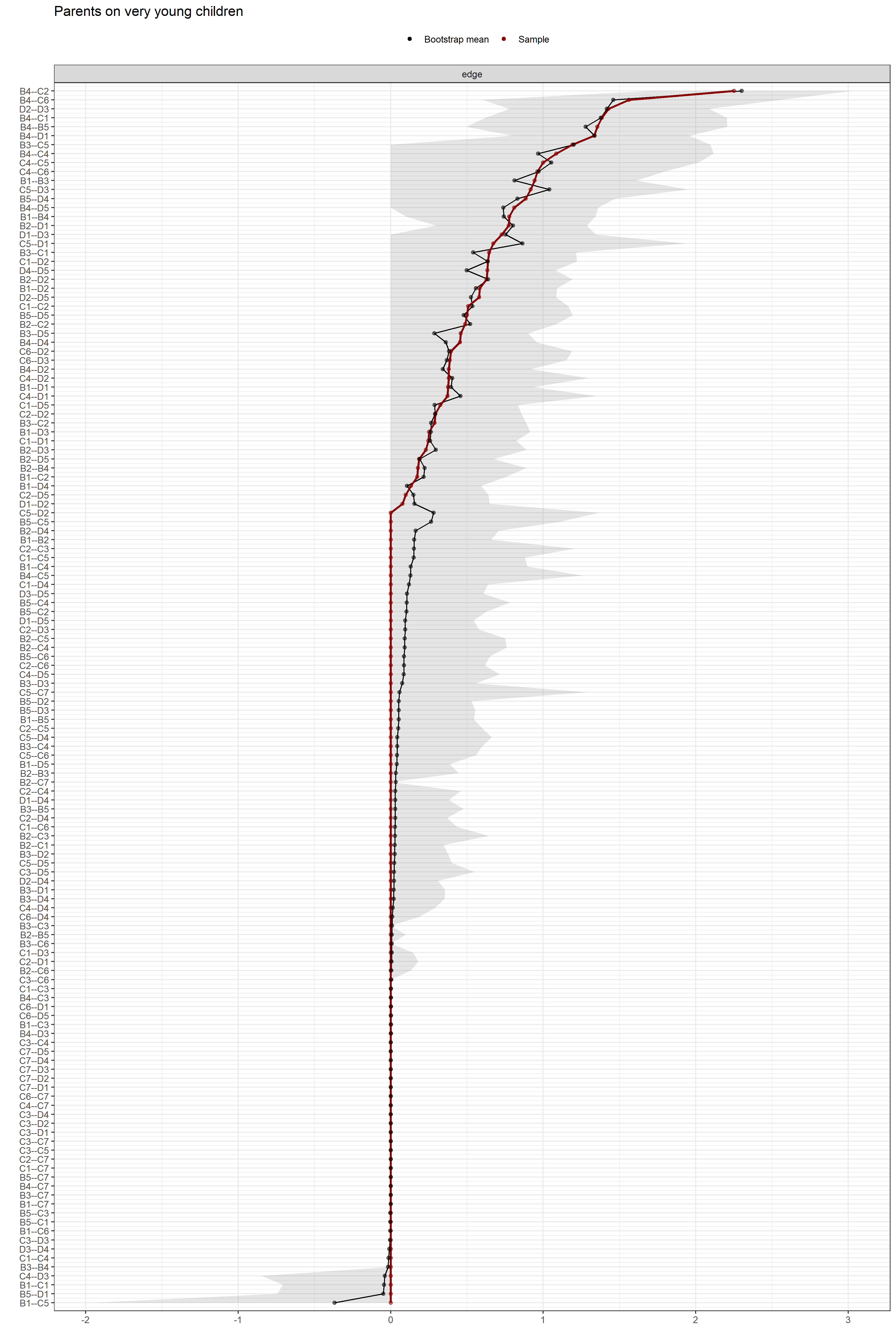

**Figure S10. Edge-weight accuracy test.** Bootstrapped confidence intervals (CIs) of the estimated edge-weights for the estimated network of the 17 DSM-IV-TR symptoms from the caregiver report of preschool-aged children dataset. The y-axis comprises all edges in the network, ordered from the edge with the highest weight-strength to the edge with the lowest weight-strength. The red dots represent the edge-weights of the network, the gray area the 95% CIs.

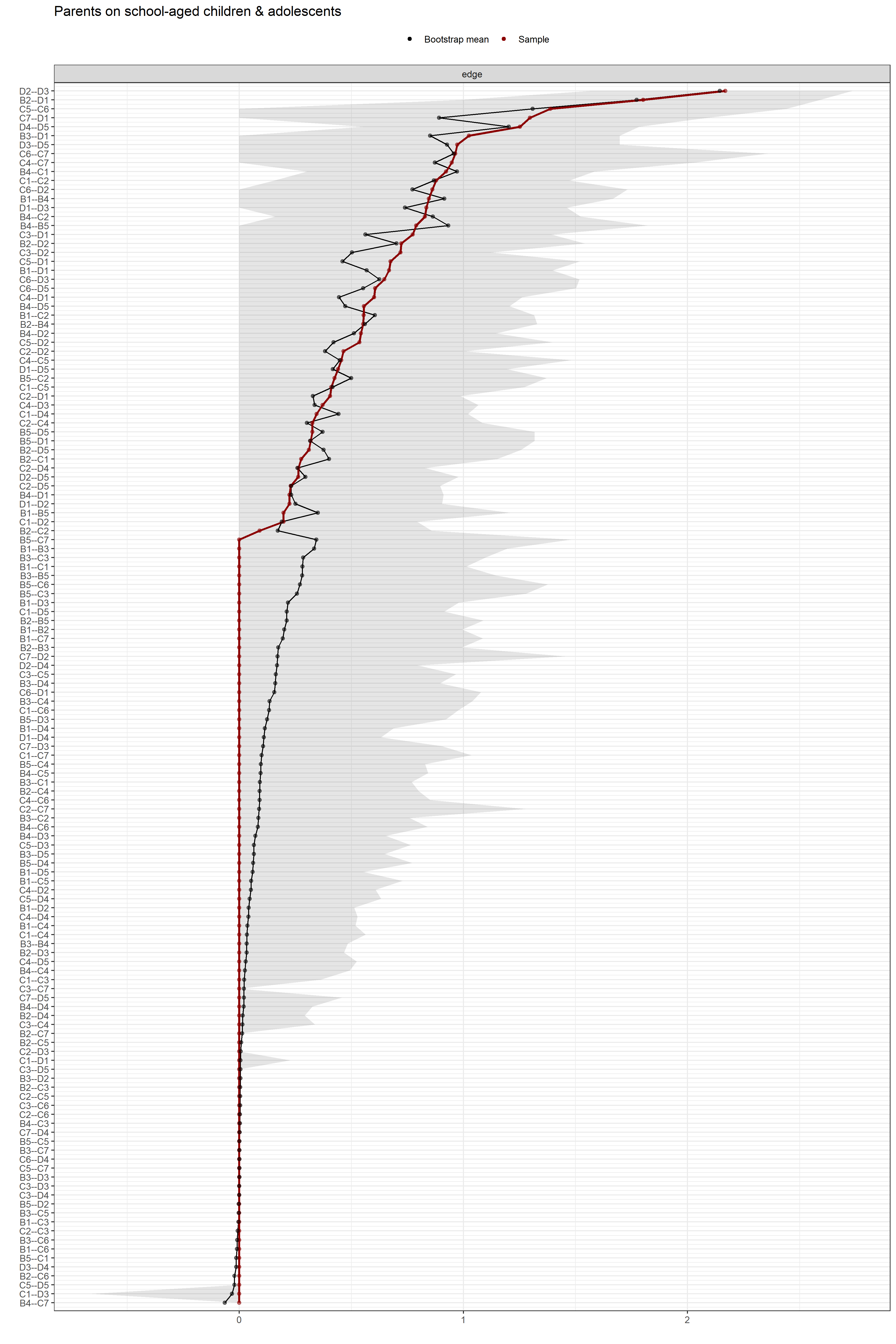

**Figure S11. Edge-weight accuracy test.** Bootstrapped confidence intervals (CIs) of the estimated edge-weights for the estimated network of the 17 DSM-IV-TR symptoms from the caregiver report of children and adolescents dataset. The y-axis comprises all edges in the network, ordered from the edge with the highest weight-strength to the edge with the lowest weight-strength. The red dots represent the edge-weights of the network, the gray area the 95% CIs.

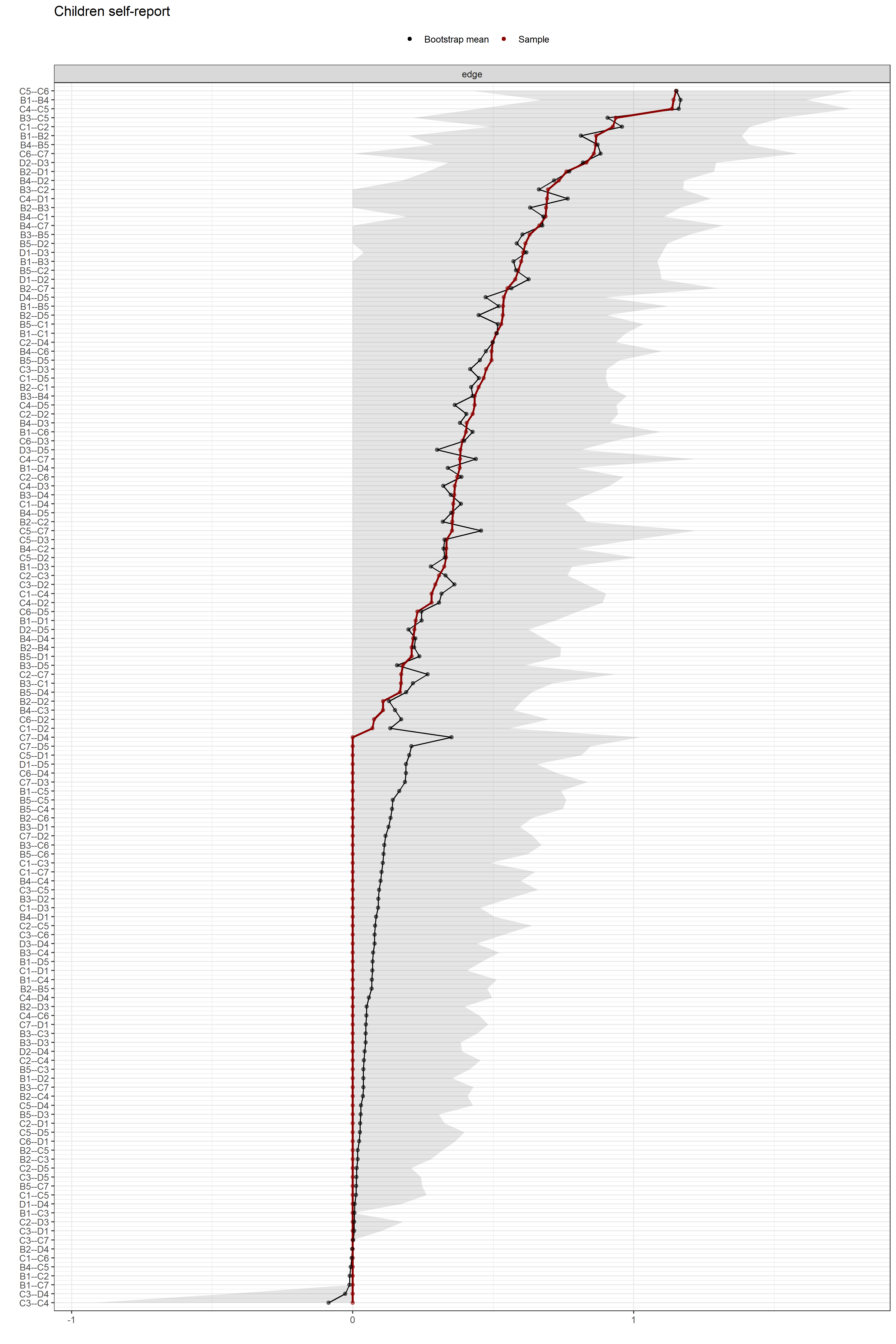

**Figure S12. Edge-weight accuracy test.** Bootstrapped confidence intervals (CIs) of the estimated edge-weights for the estimated network of the 17 DSM-IV-TR symptoms from the child self report dataset. The y-axis comprises all edges in the network, ordered from the edge with the highest weight-strength to the edge with the lowest weight-strength. The red dots represent the edge-weights of the network, the gray area the 95% CIs.

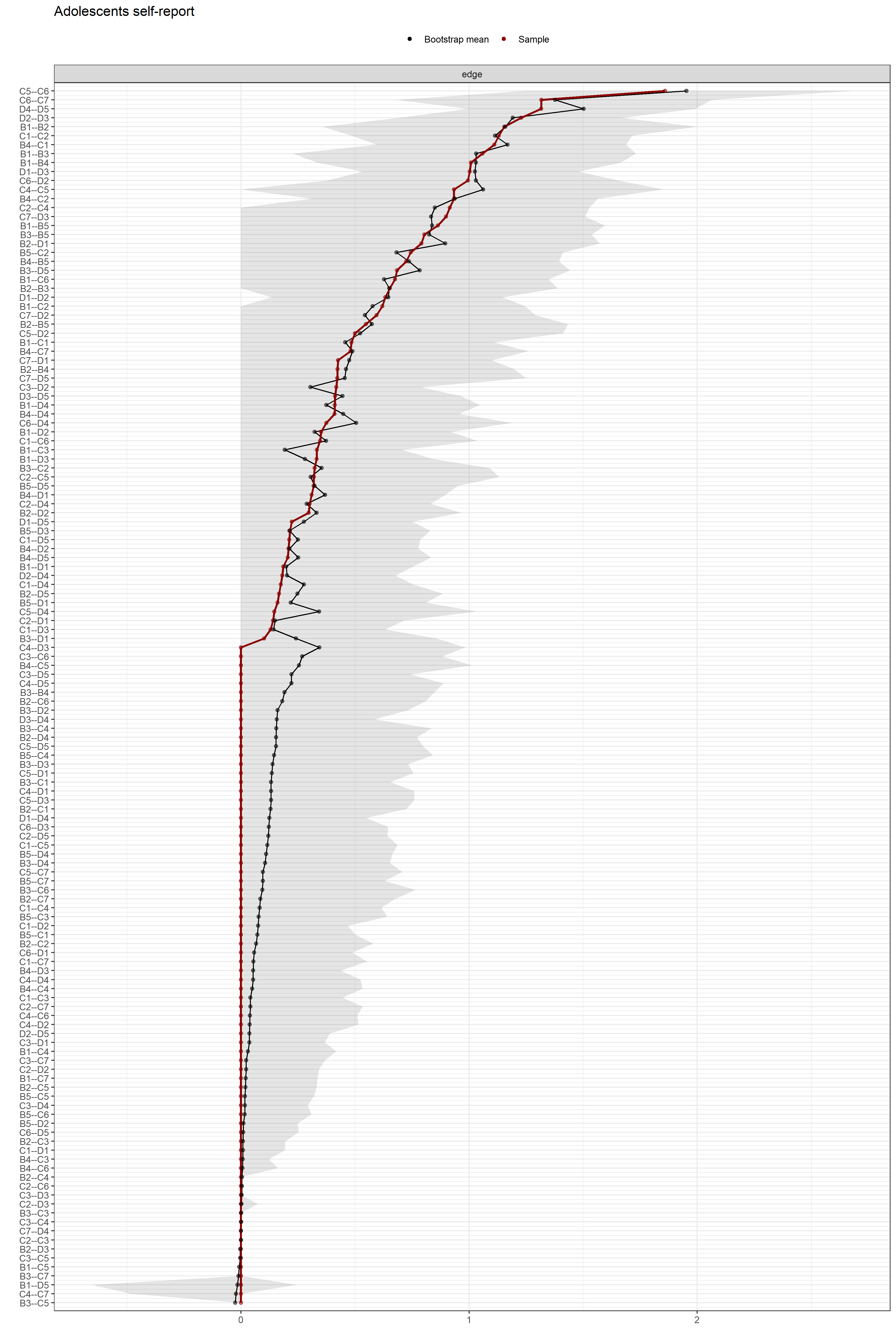

**Figure S13. Edge-weight accuracy test.** Bootstrapped confidence intervals (CIs) of the estimated edge-weights for the estimated network of the 17 DSM-IV-TR symptoms from the adolescent self-report dataset. The y-axis comprises all edges in the network, ordered from the edge with the highest weight-strength to the edge with the lowest weight-strength. The red dots represent the edge-weights of the network, the gray area the 95% CIs.

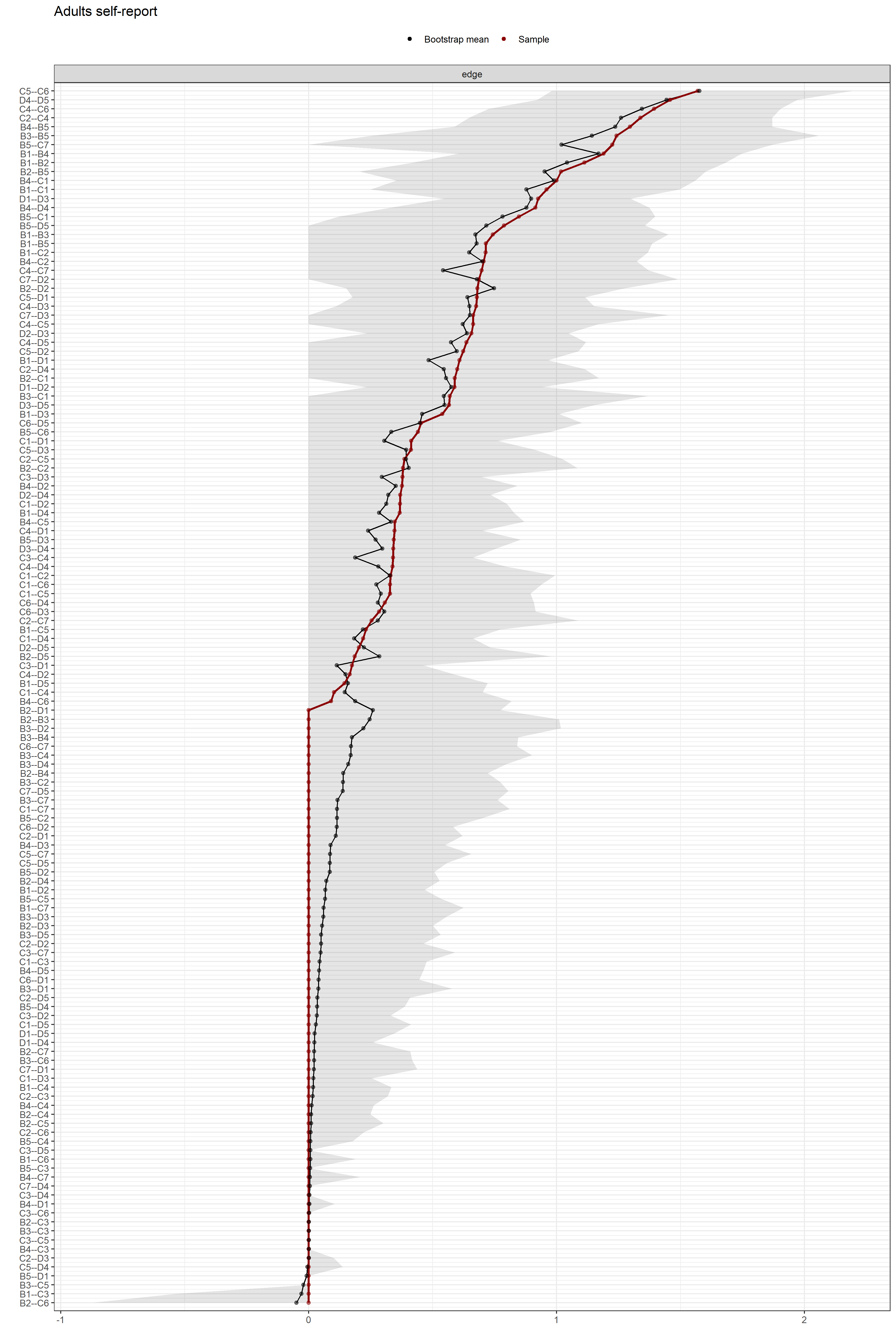

**Figure S14. Edge-weight accuracy test.** Bootstrapped confidence intervals (CIs) of the estimated edge-weights for the estimated network of the 17 DSM-IV-TR symptoms from the adult self-report dataset. The y-axis comprises all edges in the network, ordered from the edge with the highest weight-strength to the edge with the lowest weight-strength. The red dots represent the edge-weights of the network, the gray area the 95% CIs.

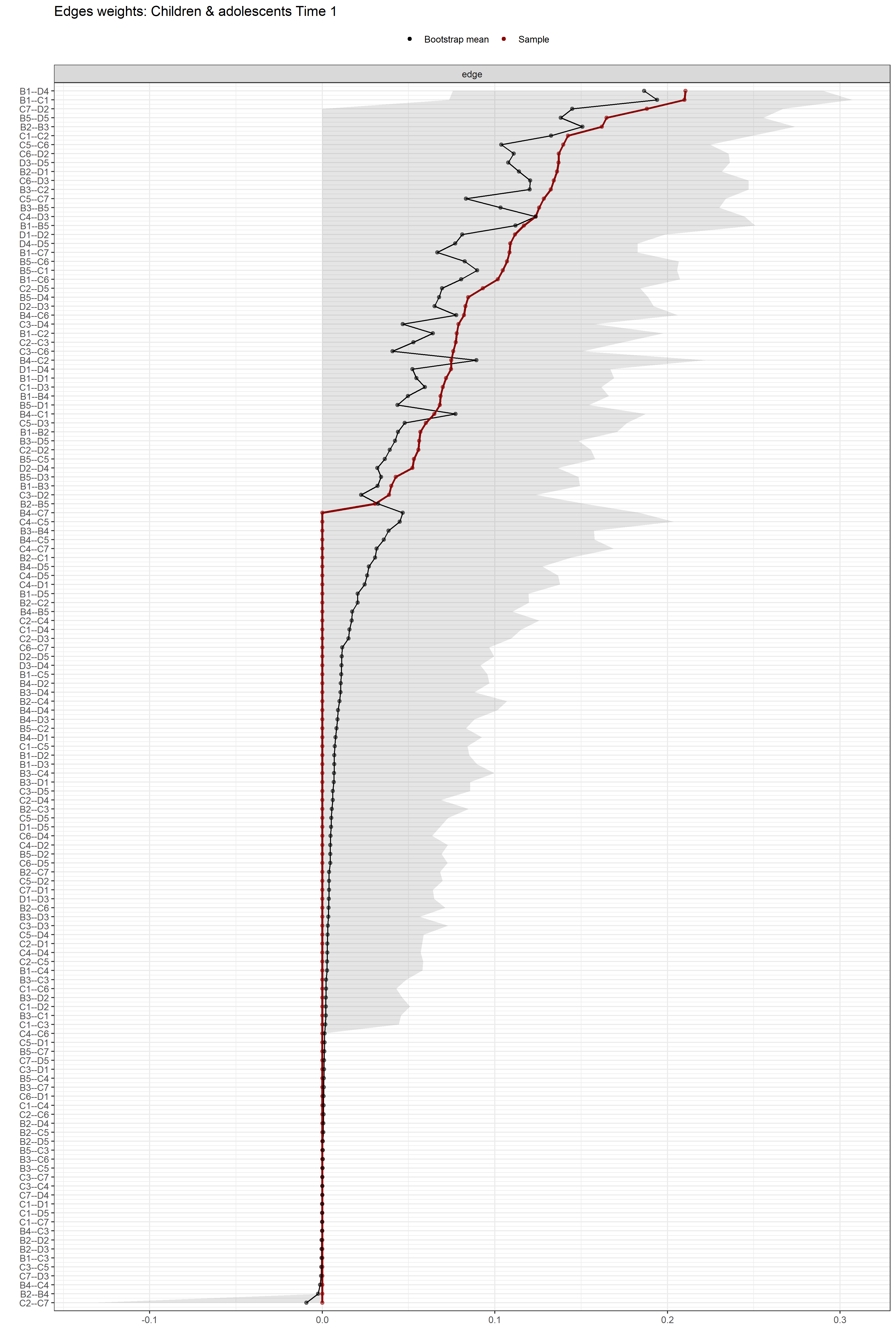

**Figure S15. Edge-weight accuracy test.** Bootstrapped confidence intervals (CIs) of the estimated edge-weights for the estimated network of the 17 DSM-IV-TR symptoms from the children and adolescents longitudinal self-report dataset (Time 1). The y-axis comprises all edges in the network, ordered from the edge with the highest weight-strength to the edge with the lowest weight-strength. The red dots represent the edge-weights of the network, the gray area the 95% CIs.

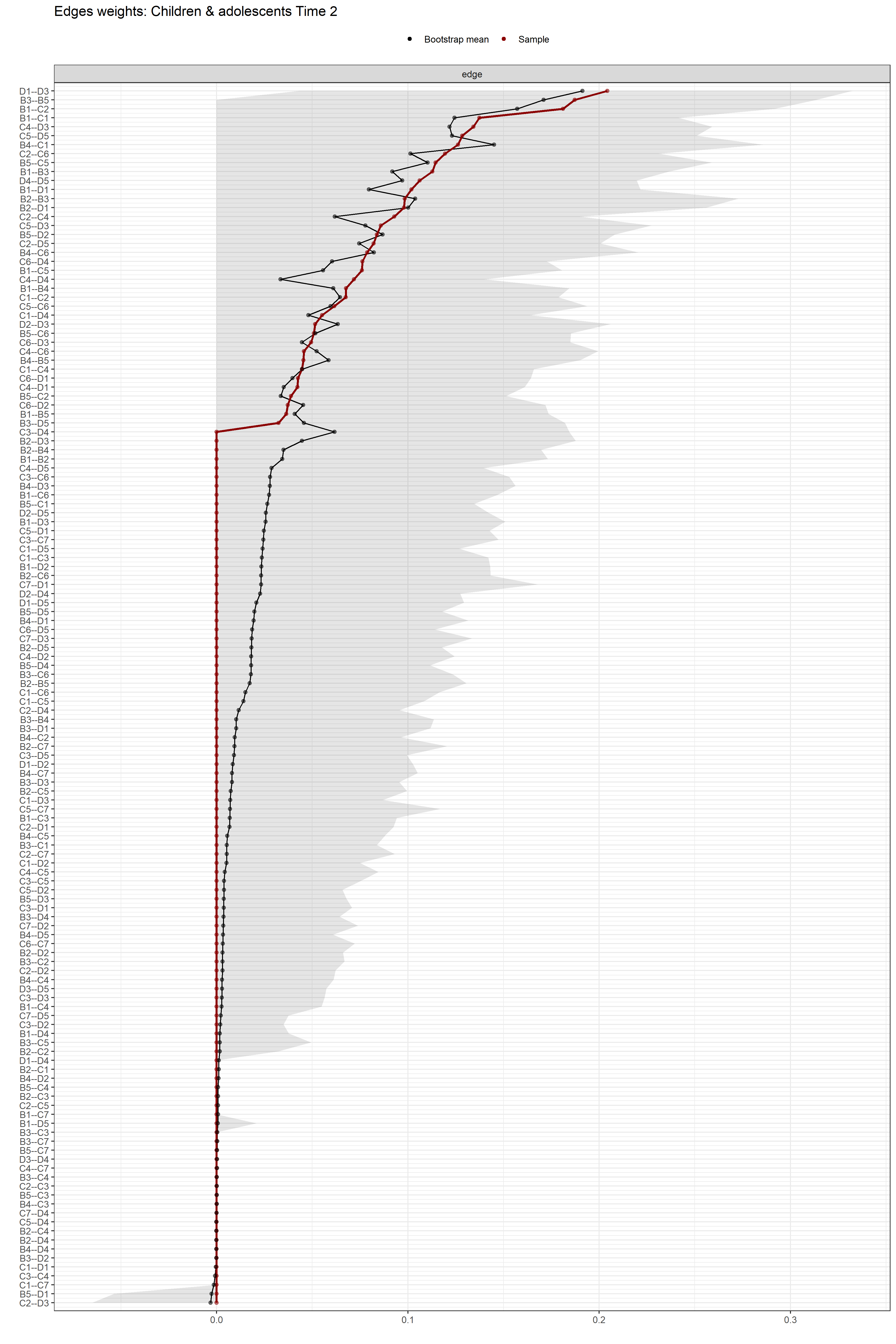

**Figure S16. Edge-weight accuracy test.** Bootstrapped confidence intervals (CIs) of the estimated edge-weights for the estimated network of the 17 DSM-IV-TR symptoms from the children and adolescents longitudinal self-report dataset (Time 2). The y-axis comprises all edges in the network, ordered from the edge with the highest weight-strength to the edge with the lowest weight-strength. The red dots represent the edge-weights of the network, the gray area the 95% CIs.

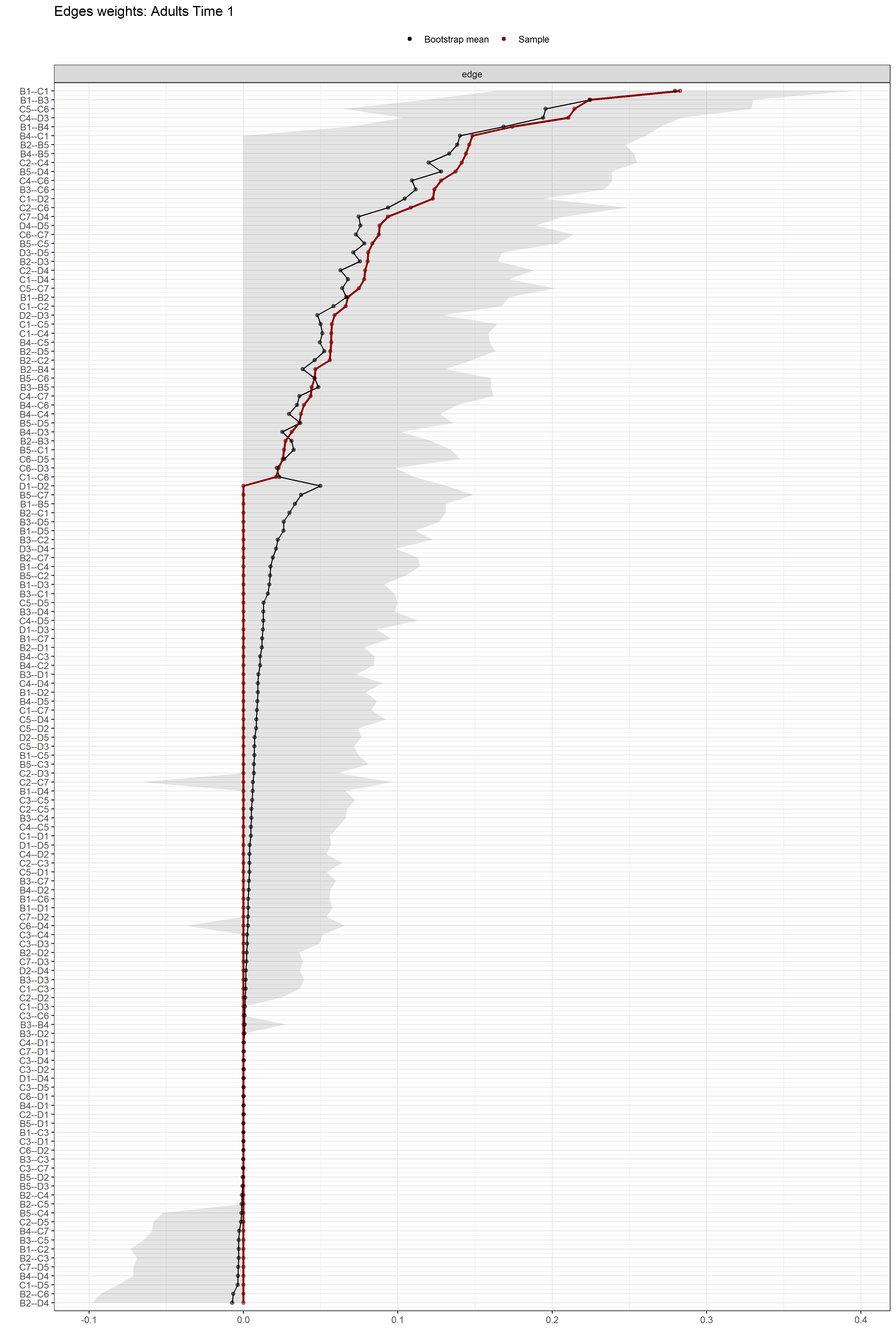

**Figure S17. Edge-weight accuracy test.** Bootstrapped confidence intervals (CIs) of the estimated edge-weights for the estimated network of the 17 DSM-IV-TR symptoms from the adult longitudinal self-report dataset (Time 1). The y-axis comprises all edges in the network, ordered from the edge with the highest weight-strength to the edge with the lowest weight-strength. The red dots represent the edge-weights of the network, the gray area the 95% CIs.

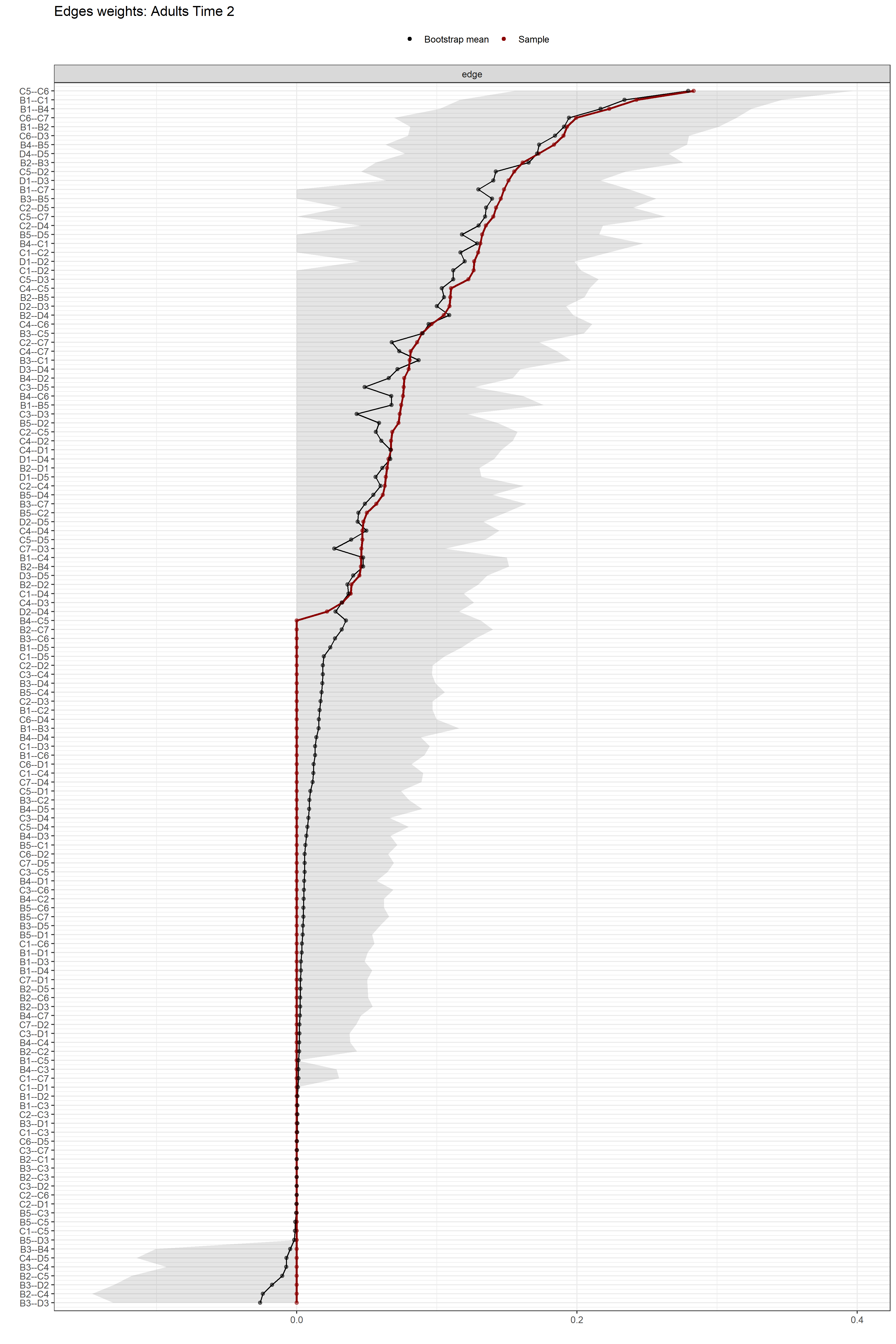

**Figure S18. Edge-weight accuracy test.** Bootstrapped confidence intervals (CIs) of the estimated edge-weights for the estimated network of the 17 DSM-IV-TR symptoms from the adult longitudinal self-report dataset (Time 2). The y-axis comprises all edges in the network, ordered from the edge with the highest weight-strength to the edge with the lowest weight-strength. The red dots represent the edge-weights of the network, the gray area the 95% CIs.
