## Supplementary materials 2 for "The Inter-Connectedness of Post-Traumatic Stress Disorder (PTSD) Symptomatology Across the Lifespan"

**Table S2.** Diagnostic algorithms

Previous studies have argued that the DSM-IV-TR criteria was insensitive for diagnosing PTSD in young children. Specifically the requirement of needing three avoidance/numbing symptoms, especially considering the capacities to verbally express symptoms (sense of a foreshortened future) or certain behavioural symptoms (loss of interest in activities) may manifest themselves differently. In response to this Scheeringa et al (2011) proposed an alternative, specifically modification to the behavioural manifestation of symptoms (for criterion D2 (irritability) is modified to also include, extreme temper tantrums. The most important and largest modification is the change in threshold for Cluster C from requiring the endorsement of 3 symptoms to 1.

|  | Traditional Diagnostic Algorithm | Alternative Algorithm |
| --- | --- | --- |
| Cluster B | Endorse one or more | Endorse one or more |
| Cluster C | Endorse three or more | Endorse one or more |
| Cluster D | Endorse two or more | Endorse two or more |

Scheeringa, M. S., Zeanah, C. H., & Cohen, J. A. (2011). PTSD in children and adolescents: toward an empirically based algorithm a. *Depression and anxiety*, *28*(9), 770-782.

### Table S3. Harmonization guide for each measure for PACT/R Measures

|  |  |  |  |  | **Checklist** | | | | | **Interview** | | | |
| --- | --- | --- | --- | --- | --- | --- | --- | --- | --- | --- | --- | --- | --- |
| ***DSM-IV language (PTSD criteria)*** | ***DSM-5 language (PTSD criteria)*** | **DSM IV**  **PTSD** | **DSM 5 PTSD** | **DSM 5 child <6** | **CATS**  **(0-1-2-3)** | **CPSS**  **(0-1-2-3)** | **CRIES-13**  **(0-1-3-5)** | **TSCC**  **(0-1-2-3)** | **UCLA PTSD RI for DSM-IV**  **(0-1-2-3-4)** | **ADIS-C**  **(0/1)** | **CAPS-CA**  **(f>=1 and i>=2))** | **IBS-A-KJ**  **(f>=1 and i>=2))** | **PTSIC**  **(0-1-2) **** |
| *recurrent and intrusive distressing recollections of the event – can be play in children* | *Recurrent, involuntary, and intrusive distressing memories of the traumatic event(s) - can be play in children;*  *In children < 6, spontaneous and intrusive memories may not appear distressing* | B1 | B1 | B1 | 1. I go over and over what happened in my mind.  OR  3. I have thoughts about what happened even when I don't want to | 1. Having upsetting thoughts or images about the event that came into your head when you didn’t want them to | 1. Do you think about it even when you don’t mean to? | 29 Can’t stop thinking abt something bad happened me  10 Remember scary things | 3. I have upsetting thoughts, pictures, or sounds of what happened come into my mind when I do not want them to. | 11. Do you have a lot of thoughts that you don’t want to have about (frightening event)? For younger children, ask: Do you ever play or draw pictures about (frightening event)? | 1Did you think about (EVENT) even when you didn’t want to? | C1. Have memories of the accident kept entering you mind? Have you ever played or drawn things that are associated with the accident? | 2. Do you ever think about [when you got hurt]? Do you think about it [when you got hurt] even when you don’t want to?  OR  3. When you close your eyes *(tester closes own eyes)*, do you ever see pictures of what happened [when you got hurt]? |
| *recurrent distressing dreams of event – can be frightening without content in children* | *recurrent distressing dreams w content or affect related to event – can be frightening without content in children* | B2 | B2 | B2 | 4. I have bad dreams about what happened | 2. Having bad dreams or nightmares | NO CORRESPONDING ITEM | 01 Bad dreams | 5. I have dreams about what happened or other bad dreams. | 12. Do you have a lot of bad dreams about (frightening event)? | 2 Did you have dreams about (EVENT)? did you have any bad dreams? | C2. Have you had bad dreams or nightmares about the accident? | 1. Some children have scary dreams about things that really happened.  Do you ever have scary dreams about [when you got hurt]? |
| *acting or feeling as if event were recurring – can be reenactment in children* | *Dissociative reactions (flashbacks) in shich the the individual feels or acts as if the traumatic event(s) were recurring. On continuum, most extreme expression = complete loss of awareness of current surroundings.*  *– can be reenactment in play in children* | B3 | B3 | B3 | NO CORRESPONDING ITEM | 3. Acting or feeling as if the event was happening again (hearing something or seeing a picture about it and feeling as if I am there again) | NO CORRESPONDING ITEM | NO CORRESPONDING ITEM | 6. I feel like I am back at the time when the bad thing happened, living through it again. | 13. Do you sometimes feel that (frightening event) is about to happen again? | 3a Did things that happen now (go on) make it seem like (EVENT) was happening all over again but it really wasn’t?  OR  3b)Did you act out things or do anything like what happened during (EVENT)? | C3a. Have you felt as if the accident was about to happen again?  **OR**  C3b. After the accident, have you done things that happened during the accident? | 8. Do you ever feel like what happened to you when you [got hurt] is happening all over again, -- even when it’s not really happening?  OR  5. Some children play games and pretend things are happening to them like when they [got hurt]. Do you ever….  OR  6. Some children play games where they pretend someone gets hurt – or pretend someone dies. Do you ever… |
| *intense psychological distress at exposure to internal or external cues that symbolize or resemble an aspect of the trauma* | *Intense or prolonged psychological distress at exposure to internal or external cues that symbolize or resemble an aspect of the traumatic event(s)* | B4 | B4 | B4 | 6. When something reminds me of what happened, I get tense and upset  OR  2. I get scared or upset when I think about what happened. | 4. Feeling upset when you think about it or hear about the event (for example, feeling scared, angry, sad, guilty, etc.) | NO CORRESPONDING ITEM | NO CORRESPONDING ITEM | 2. When something reminds me of what happened, I get very upset, afraid or sad. | 14. When things remind you of (frightening event), do you get very upset? | 4. Did you get upset (*bothered, sad, scared)* when something made you think of *(or reminded you of*) (EVENT)? | C4. Do you feel very upset when you are reminded of the accident? | NO CORRESPONDING ITEM |
| *physiological reactivity on exposure to internal or external cues that symbolize or resemble an aspect of the trauma* | *Marked physiological reactions to internal or external cues that symbolize or resemble and aspect of the traumatic event(s)*  *Child < 6:*  *Marked physiological reactions to reminders of the traumatic event(s)* | B5 | B5 | B5 | NO CORRESPONDING ITEM | 5. Having feelings in your body when you think about or hear about the event (for example, breaking out into a sweat, heart beating fast) | NO CORRESPONDING ITEM | NO CORRESPONDING ITEM | 18. When something reminds me of what happened, I have strong feelings in my body, like my heart beats fast, my head aches, or my stomach aches. | 15 When things remind you of (frightening event), do you get uncomfortable feelings in your body? For example, does your heart beat real fast, or do you sweat or shake? | 5. Did you get any feelings in your body when something made you think of (remember) (EVENT)? | C5. When you are reminded of the accident, do you sweat or tremble or does your heart beat fast? | NO CORRESPONDING ITEM |
| *efforts to avoid thoughts, feelings, or conversations assoc with the trauma* | *Avoidance or efforts to avoid distressing memories, thoughts, or feelings about or closely associated with the traumatic event(s)* | C1 | C1 | See below | 7. I try not to think about what happened. | 6. Trying not to think about, talk about, or have feelings about the event | 10. Do you try not to think about it?  OR  7. Do you try not talk about it?  OR  2. Do you try to remove it from your memory? | 39 Try not to have feelings | 9. I try not to talk about, think about, or have feelings about what happened. | 16. Do you try very hard not to think or talk about (frightening event)? | 6. Did you try not to think about [EVENT]? Did you try not to have feelings about [EVENT]? Did you try to stop the thoughts or feelings, or make them go away? | 1. D1. Have you tried not to think about the accident? Have you tried not to talk about the accident? Have you tried not to feel upset or distressed about the accident? | 2.  |
| *efforts to avoid activities, places, or people that arouse recollections of the trauma* | *Avoidance or efforts to avoid external reminders (people, places, conversations, activities, objects, situations_ that arouse distressing memories, thoughts, or feelings about or closely associated with the traumatic event(s)* | C2 | C2 | See below | 8. I try to stay away from things that remind me of what happened. | 7. Trying to avoid activities, people, or places that remind you of the traumatic event | 6. Do you stay away from reminders of it (e.g. places or situations)? | NO CORRESPONDING ITEM | 17, I try to stay away from people, places, or things that make me remember what happened. | 17. Do you try to stay away from things that remind you of (frightening event)? | 7. Did you try to stay away from people, things, or activities that make you think about *(remember)* what happened? | NO CORRESPONDING ITEM |  |
| *--* | *Child <6*  *Avoidance of or efforts to avoid activities, places, or physical reminders that arouse recollections of the traumatic event(s)* | -- | -- | C1 |  |  |  |  |  |  |  |  | 10. Some children try to stay away from scary things – because those things make them think about stuff that happened to them when they [got hurt]. Are there scary things you like to stay away from? |
| *--* | *Child <6*  *Avoidance of or efforts to avoid people, conversations, or interpersonal situations that arouse recollections of the traumatic event(s)* | -- | -- | C2 |  |  |  |  |  |  |  |  | 7. Do you ever try real hard to not think about things that happened to you when you [got hurt]? |
| *inability to recall an important aspect of the trauma* | *inability to remember an important aspect of the traumatic event(s) (typically due to dissociative amnesia and not to other factors such as head injury, alcohol, or drugs)*  *N/A for children <6* | C3 | D1 | -- | NO CORRESPONDING ITEM | 8. Not being able to remember an important part of the upsetting event | NO CORRESPONDING ITEM | 25 Forget things | 15. I have trouble remembering important parts of what happened. | cdisso05  Is there a gap in your memory of what happened during the [trauma]? | 8. Can you remember all of what happened at the time of (EVENT) or are there some parts that are hard to remember?  In this the past month, how much trouble have you had remembering important parts of what happened? | B4. (Dissociative anemesia) Did you have difficulties to recall important aspects of the accident? | 11. Some children can’t remember things that happened to them when they [got hurt]. Is it sometimes hard for you to remember things about when you [got hurt]?  OR  12. Are there bad, sad, or scary things that happened to you when you got hurt that you forgot happened? |
| *n/a* | *Persistent and exaggerated negative beliefs or expectations about oneself, others, or the world (e.g. I am bad, No one can be trusted, The world is completely dangerous. My whole nervous system is completely ruined)*  *N/A for children <6* | -- | D2 | -- | NO CORRESPONDING ITEM | NO CORRESPONDING ITEM  BUT SEE DSM5 VERSION | NO CORRESPONDING ITEM | 19 Feeling scared of men  20 Feeling scared of women  ??? | NO CORRESPONDING ITEM |  | NO CORRESPONDING ITEM  BUT SEE DSM5 VERSION |  |  |
| *n/a* | *Persistent, distorted cognitions about the cause or consequences of the traumatic event(s) that lead the individual to blame himself / herself or others.*  *N/A for children <6* | -- | D3 | -- | NO CORRESPONDING ITEM | NO CORRESPONDING ITEM  BUT SEE DSM5 VERSION | NO CORRESPONDING ITEM | *23 Feel like I did something wrong*  *???* | *14. I think that some part of what happened is my fault.* |  | NO CORRESPONDING ITEM?  SEE DSM5 VERSION  26af. Did you think that (EVENT) was your fault? [IF YES:] How was it your fault? Have you felt bad (guilty) about things you did at the time of the (EVENT)? Did you feel bad (guilty) about things you DIDN’T do at time of event? Did you think (EVENT) took place because you were bad or wrong?  27af. Did you feel bad or guilty that it was not as bad for you as it was for others? |  |  |
| *n/a* | *Persistent negative emotional state (e.g. fear, horror, anger, guilt, or shame)*  *child <6: substantially increased frequency of negative emotional states (e.g fear, guilt, sadness, shame, confusion)* | -- | D4 | C3 | NO CORRESPONDING ITEM | NO CORRESPONDING ITEM  BUT SEE DSM5 VERSION | NO CORRESPONDING ITEM | 22 Feeling stupid or bad  23 Feel like I did something wrong | *14. I think that some part of what happened is my fault.* |  | 28af. Have you felt embarrassed*, (dirty, shameful)* about what happened? Did what happened make you feel different about yourself? How? How much of the time in the past month? Has this changed or did you always feel embarrassed *(dirty, shameful*)? [IF CHANGE:] (*When did you start being* embarrassed*, [dirty, shameful*)] *Did you ever feel this way before [EVENT]* |  | 18. Some children are sad a lot.  Are you sad a lot? |
| *markedly diminished interest or participation in significant activities* | *markedly diminished interest or participation in significant activities*  *child <6: including constriction of play* | C4 | D5 | C4 | NO CORRESPONDING ITEM | 9. Having much less interest or doing things you used to do | NO CORRESPONDING ITEM | NO CORRESPONDING ITEM | 7. I feel like staying by myself and not being with my friends. |  | 9. Have you liked doing things as much as you did before the (EVENT), like doing things with friends, playing sports, or school activities? In the past month, how much of the time did you feel like you’re not able to have as much fun doing things? |  | 13. When bad things happen, some children don’t want to play as much. After you got hurt, do you not want to play as much anymore?  OR  14. Some children just don’t feel like doing fun stuff. Do you ever not feel like doing fun stuff? |
| *feeling of detachment or estrangement from others* | *feeling of detachment or estrangement from others*  *child <6:*  *socially withdrawn behavior* | C5 | D6 | C5 | NO CORRESPONDING ITEM | 10. Not feeling close to people around you | NO CORRESPONDING ITEM | 26 Feel like not in my body | 8. I feel alone inside and not close to other people. | cdisso04  When the [trauma] was happening, . Since the [trauma] happened,  feel as if your body didn’t really belong to you? / feel that you were outside your body? / feel that you weren’t really there (not really where you actually are)? | 10. Did you feel alone (all by yourself) even when other people were around? | D3. Do you feel lonely, even when other people are present? | 15. Some children feel like people don’t like them. Do you feel like people don’t like you sometimes?  OR  16. After bad things happen, some children don’t want to talk to anybody. Since you got hurt, do you not want to talk to anybody? |
| *restricted range of affect (e.g., unable to have loving feelings)* | *Persistent inability to experience positive emotions (e.g. inability to experience happiness, satisfaction, or loving feelings)*  *Child <6:*  *Persistent reduction in expression of positive emotions* | C6 | D7 | C6 | 13. I feel numb -- like I don’t have any feelings.  (NOTE – not a standard CATS item but added in many CHOP studies with the CATS author’s approval / collaboration) | 11. Not being able to have strong feelings (for example, being unable to cry or unable to feel happy) | NO CORRESPONDING ITEM | NO CORRESPONDING ITEM | 10. I have trouble feeling happiness or love.  OR  11. I have trouble feeling sadness or anger. | cdisso01  When the [trauma] was happening / Since the [trauma] happened, have you ever felt numb or empty inside? / have you ever felt so shocked that you haven’t felt anything? | 11. Were there times when it seemed like you couldn’t feel any feelings at all? | D4. Are there moments, when it seems to you, that you can cannot feel anything at all? | 17. Some children are not very happy. Is it hard for you to feel happy? |
| *sense of a foreshortened future (e.g., does not expect to have a career, marriage, children, or a normal life span)* | *n/a* | C7 | --- | -- | NO CORRESPONDING ITEM | 12. Feeling as if your future plans or hopes will not come true (for example, you will not have a job or getting married or having kids) | NO CORRESPONDING ITEM | NO CORRESPONDING ITEM | 19. I think that I wil not live a long life.  OR  21. I feel pessimistic or negative about my future. |  | 12. When you think about the future, *(growing up, getting older),* how long do you think you will live? Did you think that you would live a long time or a short time? In the past month, how much of the time have you felt that you would live a short time? |  | 19. Some children think that they will die before they grow up. Do you ever worry that you will die before you grow up? |
| *irritability or outbursts of anger* | *Irritable behavior and angry outbursts (with little or no provocation) typically expressed as verbal or physical aggression toward people or objects*  *Child <6:*  *Including extreme temper tantrums* | D2 | E1 | D1 | 9. I am grouchy or irritable | 14. Feeling irritable or having fits of anger | CRIES-13  11. Do you get easily irritable | 05 Arguing too much  11 Want yell + break things  14 Getting mad  16 Wanting to yell at people  30 Getting into fights  31 Feeling mean  40 Feeling mad | 4. I feel grouchy, angry, or mad.  OR  20. I have arguments or physical fights. | 24b. Have you had any of these problems since (frightening event)?  Losing your temper | 14. Have you been getting angry *(mad, bothered, annoyed)* more quickly than you used to? | E2. Have you felt more irritable since the accident? | 22. When some children get mad -- they scream and yell and can’t stop. Do you ever…  OR  23. Some children want to hurt other people. Do you ever…  OR  24. Some children get mad or upset real easy. Do you… |
|  | *Reckless or self-destructive behavior* | -- | E2 | -- | NO CORRESPONDING ITEM | NO CORRESPONDING ITEM  BUT SEE DSM5 VERSION | NO CORRESPONDING ITEM | 17 Wanting to hurt myself | NO CORRESPONDING ITEM |  | NO CORRESPONDING ITEM  BUT SEE DSM5 VERSION |  |  |
| *hypervigilance* | *hypervigilance* | D4 | E3 | D2 | 5. I worry that what happened will happen again | 16. Being overly careful (for example, checking to see who is around you and what is around you) | CRIES-13  12. Are you alert and watchful even when there is no obvious need to be? | 02 Afraid something bad might happen  *27 Feeling nervous*  *28 Feeling afraid* | 1. I watch out for danger or things I am afraid of. | 24d. Have you had any of these problems since (frightening event)?  Being on the “look out” a lot so you will be ready if something bad happens | 16. Have you felt like something bad was going to happen and you needed to be ready for it, sort of like watching out for danger or things that you're afraid of? | E4. Have you become more alert to danger since the accident? | 9. Do you ever feel scared to leave your home – or your room – because something bad might happen?  (better as avoidance?  OR  27. Some children are always scared something bad will happen. Are you ever…  WOULD WE INCLUDE THIS ONE? 28. Some children are always scared something bad will happen.  Are you ever…? |
| *exaggerated startle response* | *exaggerated startle response* | D5 | E4 | D3 | 10. I am jumpy and nervous | 17. Being jumpy or easily startled (for example, when someone walks up behind you) | CRIES-13  5. Do you startle more easily or feel more nervous than you did before it happened? | *13 Scared all of a sudden* | 12. I feel jumpy or startle easily, like when I hear a loud noise or when something surprises me. | 24e. Have you had any of these problems since (frightening event)?  When things happen by surprise or all of a sudden, like hearing a loud noise that you didn’t expect, does it make you “jump”? | 17. did you jump when you heard a loud noise or when something or someone surprised you? | E5. Since the accident, do you startle if you hear a loud noise or if you get surprised by something you didn’t expect? | 29. Some children get surprised real easy. Do you …?  AND  30. Some children jump up or get real scared when they hear a real loud noise.Do you ever jump up or get real scared when you hear a loud noise like this *(tester hits hand on desk or claps hands)*? |
| *difficulty concentrating* | *Problems with concentration* | D3 | E5 | D4 | 11. I have trouble keeping my mind on things | 15. Having trouble concentrating (for example, losing track of a story on the television, forgetting what you read, not paying attention in class) | CRIES-13  3. Do you have difficulties paying attention or concentrating | NO CORRESPONDING ITEM | 16. I have trouble concentrating or paying attention. | 24c. Have you had any of these problems since (frightening event)?  Having a hard time paying attention | 15. Was it hard for you to pay attention or concentrate? | E3. Have you had difficulty concentrating since the accident? | NO CORRESPONDING ITEM |
| *difficulty falling or staying asleep* | *Sleep disturbance (e.g. difficulty falling or staying asleep or restless sleep)* | D1 | E6 | D5 | 12. I sleep poorly | 13. Having trouble falling or staying asleep | CRIES-13  13. Do you have sleep problems? | NO CORRESPONDING ITEM | 13. I have trouble going to sleep or I wake up often during the night. | 24a. Have you had any of these problems since (frightening event)?  Trouble sleeping | 13. did you have trouble sleeping? | E1. Have you had trouble sleeping since the accident? | 20. Some children can’t fall asleep when they really want to. Is it ever hard for you to fall asleep when you really want to?  OR  21. Some children can’t fall asleep because they are thinking about things that happened.  Is it ever hard for you to fall asleep – because you are thinking about things that happened when you got hurt? |

** *Format for all PTSIC items: Some children [XXX]. Do you ever [XXX]. [IF YES] Do you [XXX] a real lot – like almost every day? Or, do you just [XXX] sometimes?: Scored as No = 0; Sometimes = 1; A real lot / Almost every day = 2.*

**Figure S19**. Caregiver Report Sample Selection Process

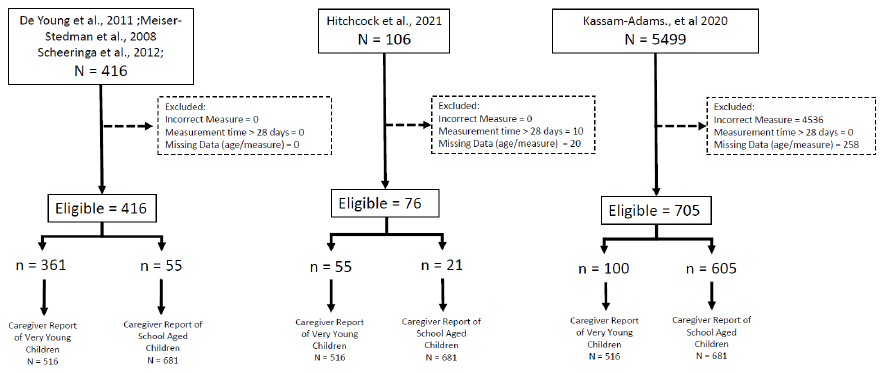

Excluded:

Incorrect Measure: 4536

Measurement time < 28 = 0

Missing Data (age/measure) = 205

Excluded:

Incorrect Measure: 0

Measurement time < 28 = 10

Missing Data (age/measure) = 20

Excluded:

Incorrect Measure: 0

Measurement time < 28 = 0

Missing Data (age/measure) = 0

758 eligible

n = 654

n = 104

76 eligible

n = 21

n = 55

n = 55

n = 361

416 eligible

Caregiver-Report of

Preschool-Aged Children

N = 520

Caregiver-Report of

School-Aged Children

N = 730

Caregiver-Report of

School-Aged Children

N = 730

Caregiver-Report of

Preschool-Aged Children

N = 520

Caregiver-Report of

Preschool-Aged Children

N = 520

Caregiver-Report of

School-Aged Children

N = 730

Kassam-Adamns et al., 2020

N = 5499

Hitchcock et al., 2021

N = 106

De Young et al., 2011

Meiser-Stedman et al., 2008

Scheeringa et al., 2012

N = 416

**Figure S20**. Self-Report Sample Selection Process

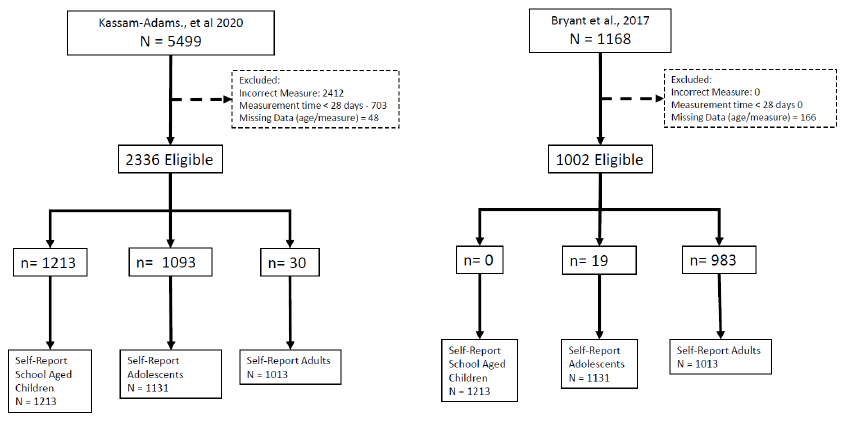

Kassam-Adams et al., 2020

N = 5499

Bryant et al., 2017

N = 1168

Excluded:

Incorrect Measure: 0

Measurement time < 28 days = 0

Missing Data (age/measure) = 184

Excluded:

Incorrect Measure: 2412

Measurement time < 28 = 703

Missing Data (age/measure) = 316

2068 eligible

984 eligible

n= 19

n= 0

n= 965

Self-Report

School Aged Children

N = 1079

Self-Report

Adolescents

N = 978

Self-Report

Adults

N = 995

n= 30

Self-Report

Adults

N = 995

n= 959

Self-Report

School Aged Children

N = 1079

Self-Report

Adolescents

N = 978

n= 1079

**Figure S21**. Longitudinal Data Sample Selection Process

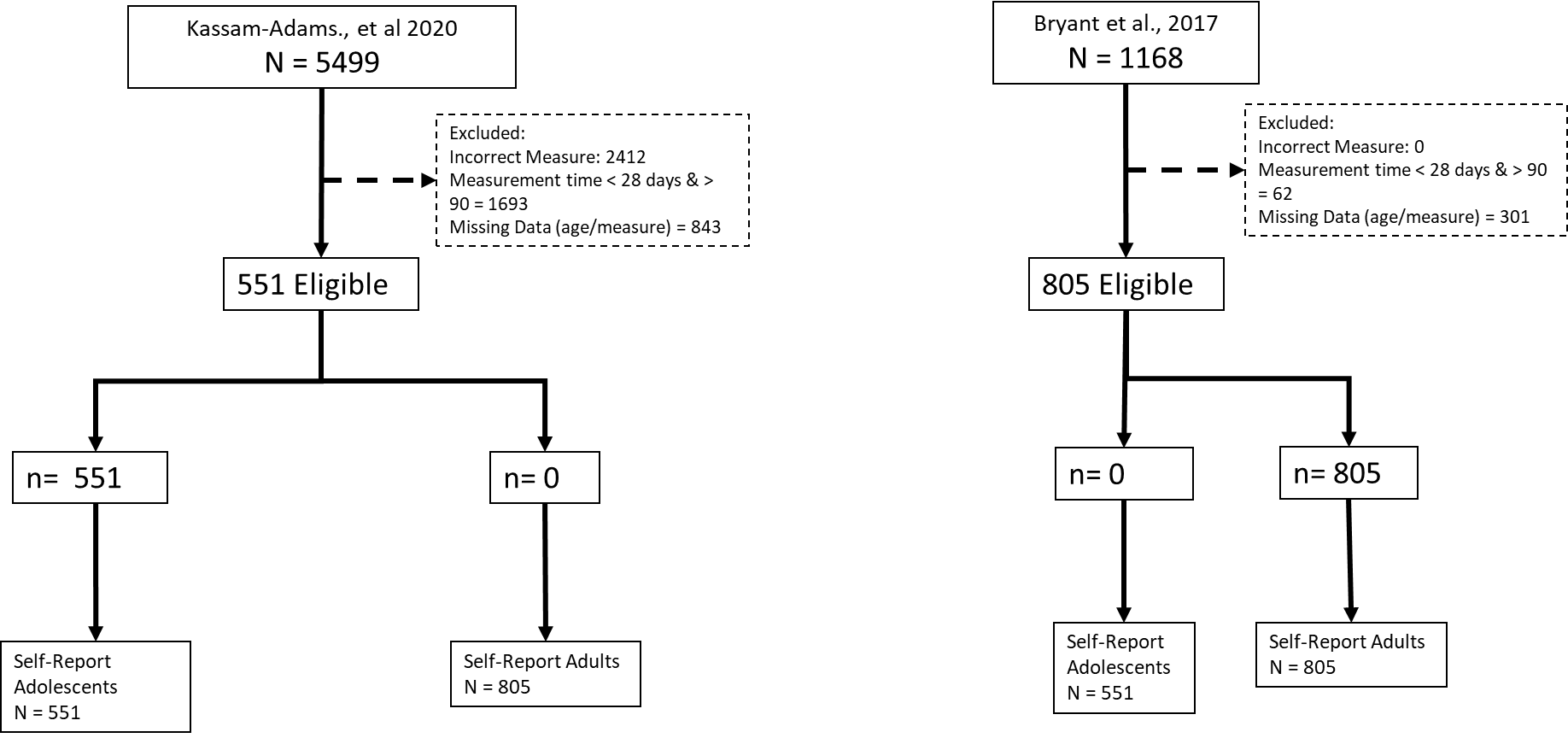

n= 0

809 Eligibile

n= 0

n= 0

Excluded:

Incorrect Measure: 2604

Measurement time < 28 days & > 90 = 1693

Missing Data (age/measure) = 843

Kassam-Adams et al., 2020

N = 5499

Bryant et al., 2017

N = 1168

Excluded:

Incorrect Measure: 0

Measurement time < 28 days & > 90 = 62

Missing Data (age/measure) = 297

n= 809

Self-Report

Adults

N = 809

Self-Report

Adults

N = 809

Self-Report

Adolescents

N = 359

Self-Report

Adolescents

N = 359

359 Eligibile

n= 359
